# The quest for the best: manual, atlas- and spatial prior-based delineation of locus coeruleus

**DOI:** 10.1101/2025.11.24.25340859

**Authors:** Olga Dmitrichenko, Giuseppina Baldizzi, Tatjana Schmidt, Aurélie Bussy, Olivier Colliot, Antoine Lutti, Ferath Kherif, Bogdan Draganski

## Abstract

Despite advances in neuromelanin-sensitive brain imaging and a plethora of manual labelling, atlas- and signal intensity-based software solutions, the reliable non-invasive delineation of the locus coeruleus (LC) in the human brain stem remains challenging. We sought to evaluate the spatial accuracy and consistency of atlas-and probabilistic spatial prior-based LC delineation. We acquired neuromelanin-sensitive magnetic resonance imaging data in healthy volunteers (n = 24; mean age 40.0 ± 16.8 years; 42% females). Manual labelling by 9 raters performed twice provided the basis for individual- and group-level comparisons with the automated delineation methods. For the atlas-based labelling, we separately tested seven open-access LC atlases, the averaged output of the manual labelling, and a consensus reference representing the atlases’ overlap. Each one of the atlases served as spatial prior for automated LC delineation in a probabilistic segmentation framework. Manual labelling showed moderate inter-rater agreement (mean Dice = 0.7), with higher delineation variability in the rostral and caudal LC. For the atlas-based labelling we observed a low spatial concordance for all open-access atlases (Dice = 0.2-0.4) with inconsistent boundary accuracy and volume similarity indices relative to the consensus reference. In the same comparison, the averaged manual labelling atlas showed higher spatial overlap (Dice = 0.6). Probabilistic delineation using spatial priors showed the strongest voxel-wise similarity with manual labelling (correlation coefficient r = 0.3) when the averaged manual labelling atlas was used as prior. Principal component analysis confirmed the greater spatial compactness for atlas-based labelling in comparison with spatial prior-based delineation, underscoring method-dependent differences in anatomical organization. Our results highlight the potential advantage of atlas-based labelling for robust and spatially reliable LC identification. The observed variability across methods and atlases calls for harmonised validation strategies and context-sensitive approaches that improve reliability.

## 1 Introduction

The locus coeruleus (LC), referred to as the “blue spot” due to its high neuromelanin content, is a small brainstem nucleus measuring approximately 15mm rostro-caudally and 2.5mm medio-laterally (Baker et al., 1989; Fernandes et al., 2012; Mouton et al., 1994; Pamphlett et al., 2018, 2020). Given LCs critical involvement in neurodegenerative disorders, there is a plenitude of studies providing evidence for disease-related anatomical differences (Lakhani et al., 2024; Trujillo et al., 2023; Zecca et al., 2002; Zecca, Youdim, et al., 2004). Consequently, there is a growing interest in establishing LC-centred imaging biomarkers, sensitive to early presymptomatic stages in neurodegeneration and especially in Alzheimer’s disease (Braak et al., 2011; Satoh & Iijima, 2019).

Unlike the basal ganglia, the LC contains noradrenergic neurons with relatively low iron and high copper content (Zecca, Stroppolo, et al., 2004), which makes it challenging to visualise *in vivo* using conventional iron-sensitive magnetic resonance imaging (MRI). Dedicated neuromelanin-sensitive MRI (NM-MRI) protocols improve LCs visualisation by leveraging the specific paramagnetic properties of neuromelanin (Liu et al., 2017) that correlate with post-mortem LC localisation (Sasaki et al., 2006). NM-MRI has prompted a line of research on the delineation of the LC using the absolute and relative signal intensity characteristics, or combined with LC-specific spatial priors (Flores et al., 2025) for inferences about disease-specific effects (Lee et al., 2024). This renders NM-MRIs well-suited for LCs manual labelling in small-sized group studies (Clewett et al., 2016; Mather & Harley, 2016), but less practical for large-scale cohorts.

Atlas-based labelling of the LC represents a widely used alternative to the manual delineation. However, the accuracy of atlas-led LC definition largely depends on the atlas representativeness with reference to demographics, sample recruitment and sample size characteristics (Lee et al., 2024; Manaye et al., 1995). This could explain the modest spatial overlap of several freely available LC atlases (Dahl et al., 2019; Liu et al., 2019; Tona et al., 2017; Ye et al., 2021). Another source of potential delineation accuracy loss is the spatial registration precision between standard and individuals’ native space. The anisotropic resolution of the majority of NM-MRI protocols, the reduced field-of-view acquisition, and inferior–superior positioning rather than alignment to the anterior commissure–posterior commissure line, covering only the brainstem, introduce additional challenges for spatial registration (Trujillo et al., 2023). Transforming individual LC atlases into standard space typically results in moderate cross-subject overlap (<62%), with variability further influenced by image quality and sample characteristics (Yi et al., 2023). This is supported by empirical evidence showing that smaller structures are disproportionally more impacted by the choice of the applied linear and non-linear interpolations of spatial registration (Ashburner & Friston, 2011; Avants et al., 2008; Klein et al., 2009). Advances in diffeomorphic registration algorithms focusing on the brainstem carry the promise of mitigating this issue (Ashburner, 2007; Avants et al., 2008; Betts et al., 2017; Fonov et al., 2011).

Advanced computational anatomy techniques using MRI and evolving feature extraction algorithms overcome the limitations of manual and atlas-based labelling methods (Ashburner et al., 2003). The recent developments in atlas-guided or machine!ZIlearning-driven feature extraction (Aganj et al., 2024; Dünnwald et al., 2021) offer major benefits in terms of time efficiency and consistency. However, the LC’s fusiform elongated shape and anatomical location make the robust and reliable delineation difficult (Liu et al., 2017; Yi et al., 2023). Automated approaches such as the funnel-tip method successfully benchmarked against manual labelling (intraclass correlation coefficient = 0.91) (Sibahi et al., 2023).

Considering the lack of a non-invasive in vivo imaging technique that provides the ground truth for LC spatial extent, the existing literature encompasses a wide range of arbitrarily chosen methods for validating the reported delineation accuracy. To address this challenge, we provide empirical evidence guiding to informed decision about the choice of a particular LC atlas by quantifying spatial consistency and alignment accuracy. Here, we systematically evaluated three commonly used strategies for LC delineation: manual labelling, atlas-based, and LC-specific spatial prior–based delineation. We use multi-contrast MRI acquisitions with repeated annotations from several expert raters and comparisons across existing LC atlases, that allows assessment of spatial variability, as well as reproducibility. This approach allows for differentiating between methodological and anatomical sources of variation. While deep-learning approaches have recently shown promise for automated LC segmentation, they generally rely on large, heterogeneous datasets and can be difficult to interpret in terms of anatomical features. We propose a complementary approach to build a consensus LC atlas within established frameworks that capture inter-individual variability. The consensus altas can then be used for future methods (including deep learning) and clinical studies.

## 2 Methods

### 2.1 Participants

For this study, we recruited 26 healthy volunteers. Two participants were excluded from further analysis because of inconsistent NM-MRI protocol parameters, resulting in a final sample of 24 individuals (10 females, 14 males; mean age 40.0 ± 16.8 years). The study was approved by the Institutional Ethics Committee of the University of Lausanne and informed written consent was obtained from all participants (CER-VD, project number PB_2018-00038 (239/09)).

### 2.2 MRI data acquisition

All MRI data were acquired on a 3T whole-body system (Prisma, Siemens Healthcare, Erlangen Germany) with a standard 32-channel radiofrequency receive head coil and body coil for transmission.

We used a T1-weighted (T1w) magnetization-prepared rapid acquisition gradient echo (MPRAGE) 3D gradient-recalled inversion recovery (GR-IR) protocol with the following parameters: repetition time (TR) = 2000 ms, echo time (TE) = 2.39 ms, inversion time (TI) = 920 ms, and flip angle α = 9° yielding isotropic voxel sizes of 1×1×1 mm.

The neuromelanin-sensitive protocol consisted of 2D gradient-echo (GRE) acquisitions in transverse orientation with enabled magnetization transfer contrast, optimized for neuromelanin with TR = 274ms, TE = 2.28ms, flip angle = 40°, voxel size = 0.51×0.51×3.24mm, and field of view (FoV) = 180mm providing 12 interleaved slices with five signal averages.

### 2.3 Unified approach and spatial registration estimates

We evaluated three complementary approaches for LC delineation: atlas-based labelling and SPM12s “unified segmentation” (Wellcome Centre for Human Neuroimaging, University College London, UK) augmented with a LC-specific tissue prior whilst comparing with manual labelling by trained raters. For each approach, analyses were repeated across several freely available LC atlases to independently assess the effects of atlas choice versus delineation method.

Aiming to evaluate the different methods’ output in both native and standard Montreal Neurological Institute (MNI) space, we estimated individuals’ spatial registration parameters using whole-brain T1w data, which provides the deformation fields and serves as anatomical reference to anchor the reduced field-of-view of NM-MRI scans. To this end, we applied SPM12s unified segmentation with enhanced tissue priors (Lorio et al., 2016), followed by registration using the geodesic shooting algorithm. To ensure spatial consistency in the NM-MRI data, we linearly registered the NM-MRI to the native space T1w images using 6-parameter rigid-body registration with nearest neighbour interpolation. During this step, we resampled images from their original voxel size (0.51×0.51×3.24mm) to an isotropic resolution of 1×1×1mm (full preprocessing in **Fig. 1, A-B).**

**Figure 1.**
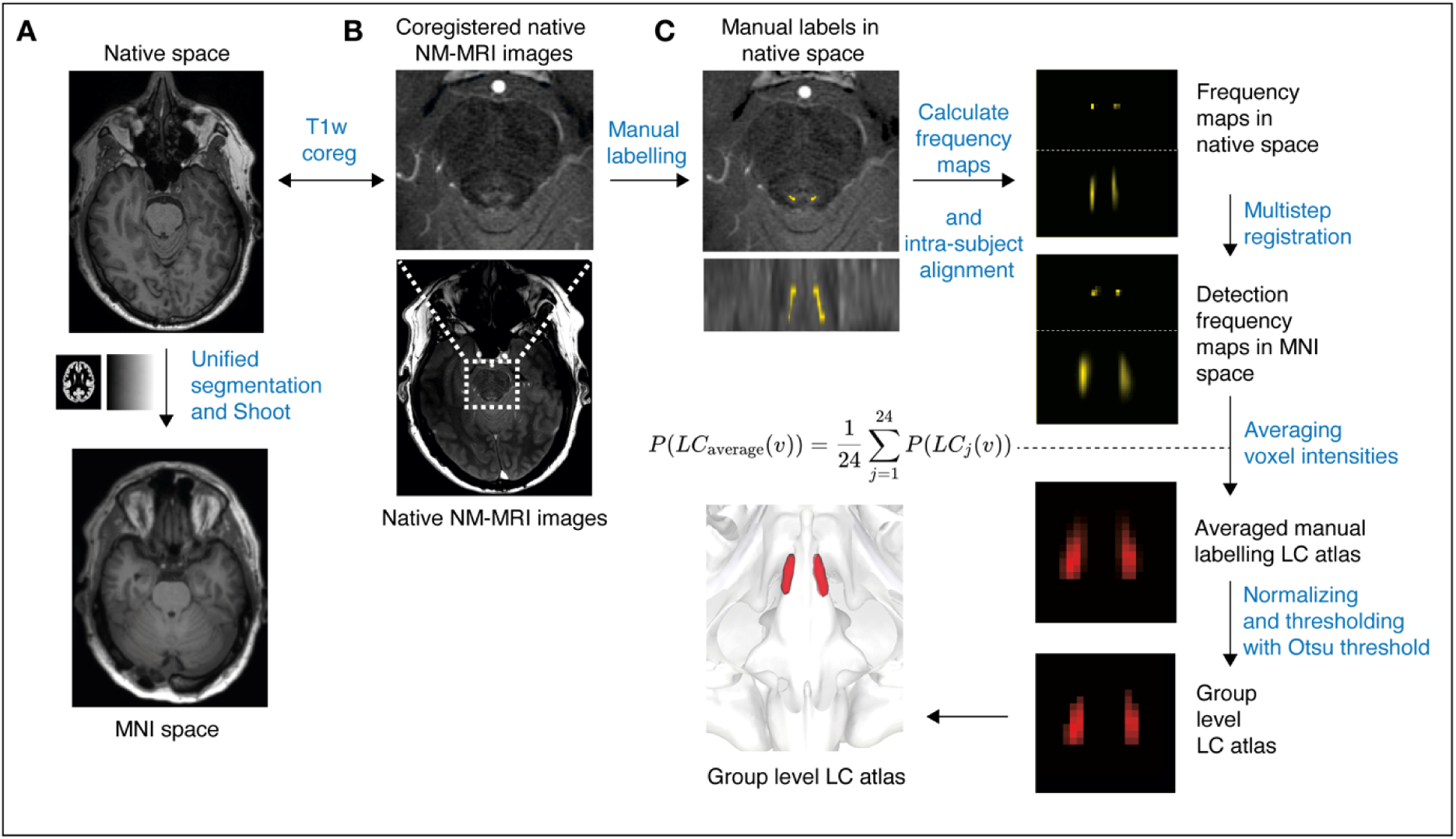
**A.** T1w images segmented and spatially registered to MNI space. **B.** NM co-registered with native space T1w images and resampled to isotropic resolution. **C.** Manual labels in native space NM-MRI space aligned to T1w images, converted to detection frequency maps, and registered to MNI space. Detection frequencies maps averaged, spatially registered, and thresholded to create a group-level LC atlas. *Abbreviations: T1w = T1-weighted; MNI = Montreal Neurological Institute; NM-MRI = neuromelanin-sensitive magnetic resonance imaging; LC = locus coeruleus*.

### 2.4 Manual labelling

The LC was manually labelled on 24 NM-MRIs by 9 raters twice in all three planes, following instructions on software use and LC identification. Raters were not blinded to subject identifiers but completed all first pass labelling prior to initiating the second round. For visualisation and labelling, we used the MINC Toolkit (version 1.9.18, https://bic-mni.github.io).

We assessed the intra-rater reliability of manual labelling using the Dice-Sørensen coefficient (DSC) for each annotated voxel. The DSC was calculated as twice the number of voxels that were labelled in both annotations (from the 2 repetitions of the same rater), divided by the sum of the voxels labelled in each annotation. Inter-rater reliability was assessed using the Fleiss’ Kappa index. Since the LC contains only a few voxels, we restricted the search volume to a region of interest (ROI) surrounding the expected LC location, defined by its coordinates in MNI space (x=82–98, y=80–100, z=0–60) to reduce potential biases from non-LC voxels. The Fleiss’ Kappa index was calculated on a voxel basis within the ROI across all raters and datasets.

We created detection frequency maps by accumulating labels at the voxel level across raters and repetitions, reflecting raw counts. These maps were then co-registered and resampled to 1×1×1mm with trilinear interpolation, followed by warping in MNI space using the already estimated diffeomorphic spatial registration parameters. We generated a group-level atlas by averaging detection frequencies across subjects (from now on denoted as averaged manual labelling atlas or group-level atlas). Using Otsu’s thresholding, we excluded low-agreement voxels and minimized alterations from the spatial registration in the averaged manual labelling atlas. The remaining values were either scaled to the [0, 1] range for group-level atlas creation or set to 1 for a binary version (see Suppl. Fig. 1). At each step, we visually inspected image quality and spatial alignment (see **Fig. 1, C).**

### 2.5 Overview of included LC atlases

The main characteristics of the open-access LC atlases (Betts et al., 2019; Dahl et al., 2019; Keren et al., 2009; Lee et al., 2024; Liu et al., 2017; Tona et al., 2017; Ye et al., 2021) included in our analyses are summarized in **Table 1**.

**Table 1.**
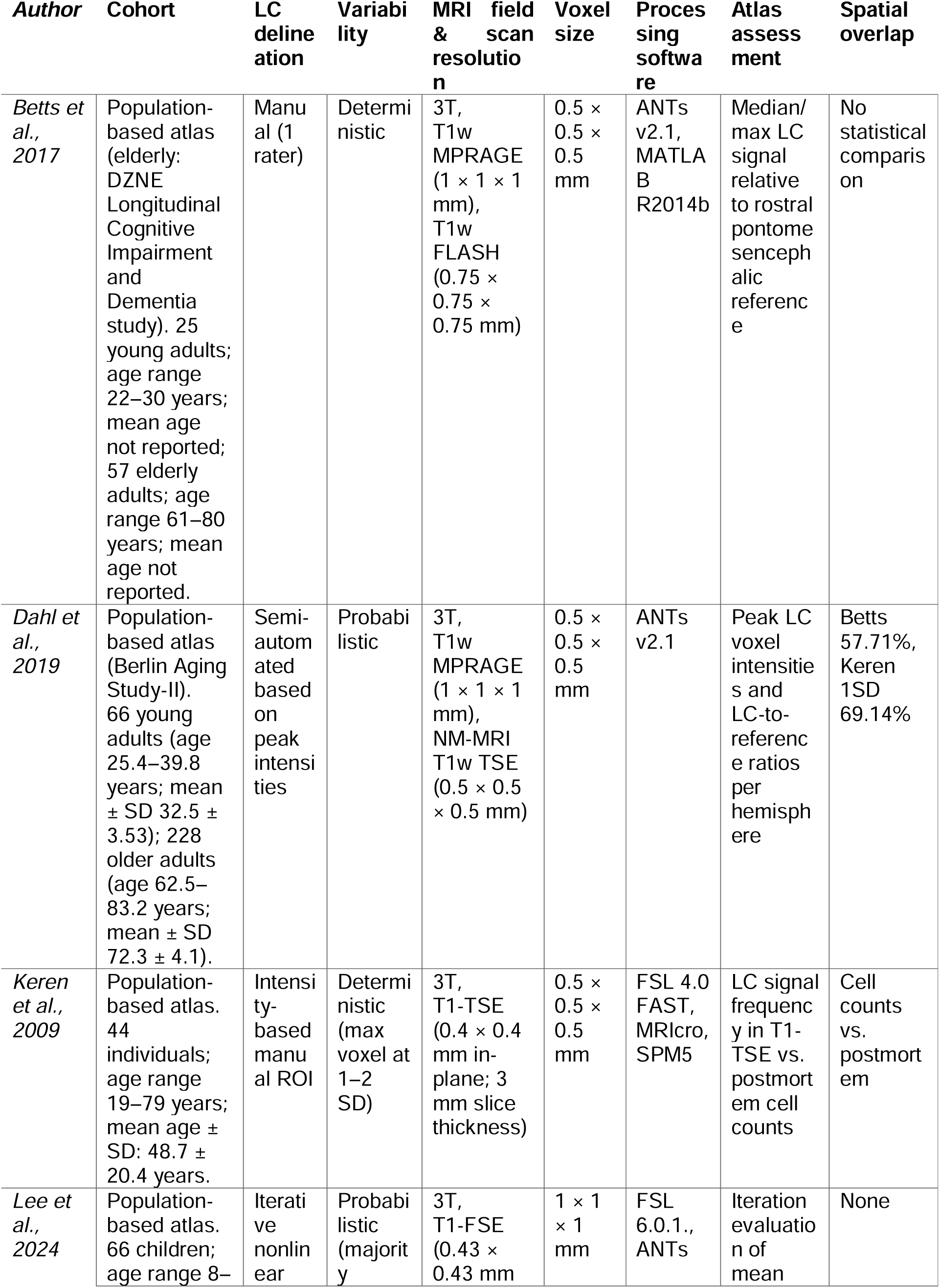
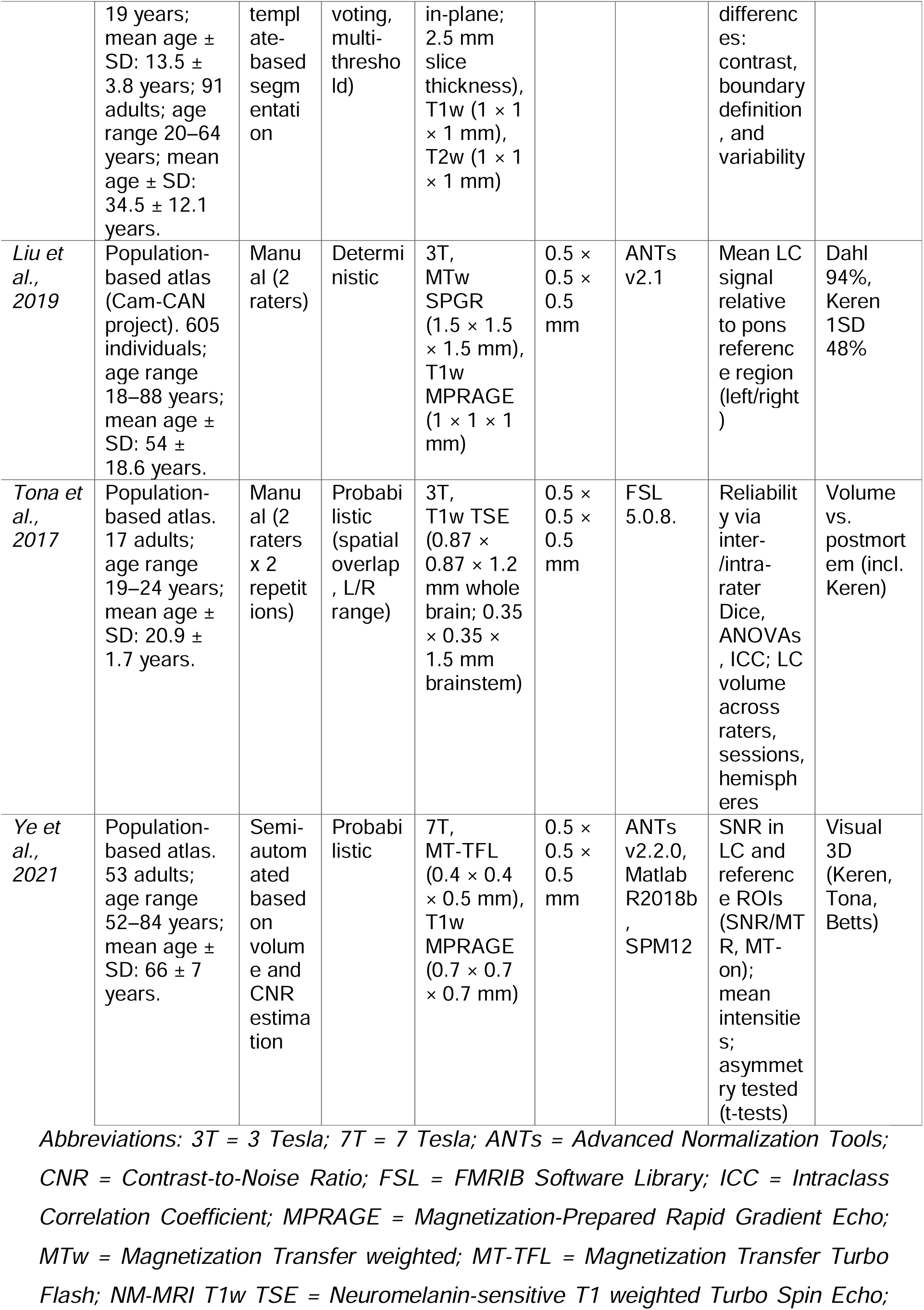
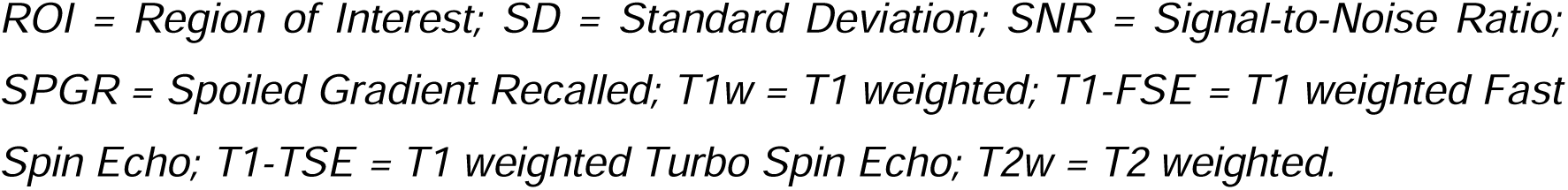
Overview of included LC atlases.

### 2.6 Atlas-based labelling

To enable a fair comparison, we used two atlas sets: the freely available LC atlases and the group-level LC atlas generated by averaging manual labels (Suppl. Table 1B). For each atlas, we generated probabilistic and maximum probability labels. We integrated those labels alongside the 125 cortical and subcortical labels from the “MICCAI 2012 Grand Challenge and Workshop on Multi-Atlas Labeling” (https://my.vanderbilt.edu/masi/about-us/resources-data). To achieve this, we resampled the atlases to 1.5×1.5×1.5mm to simplify integration into the MICCAI label set. The LC label was excluded from the brainstem label (see **Fig. 2**). The atlas-based labelling included spatial registration and labelling in individuals’ native space (Ashburner & Friston, 2011).

**Figure 2.**
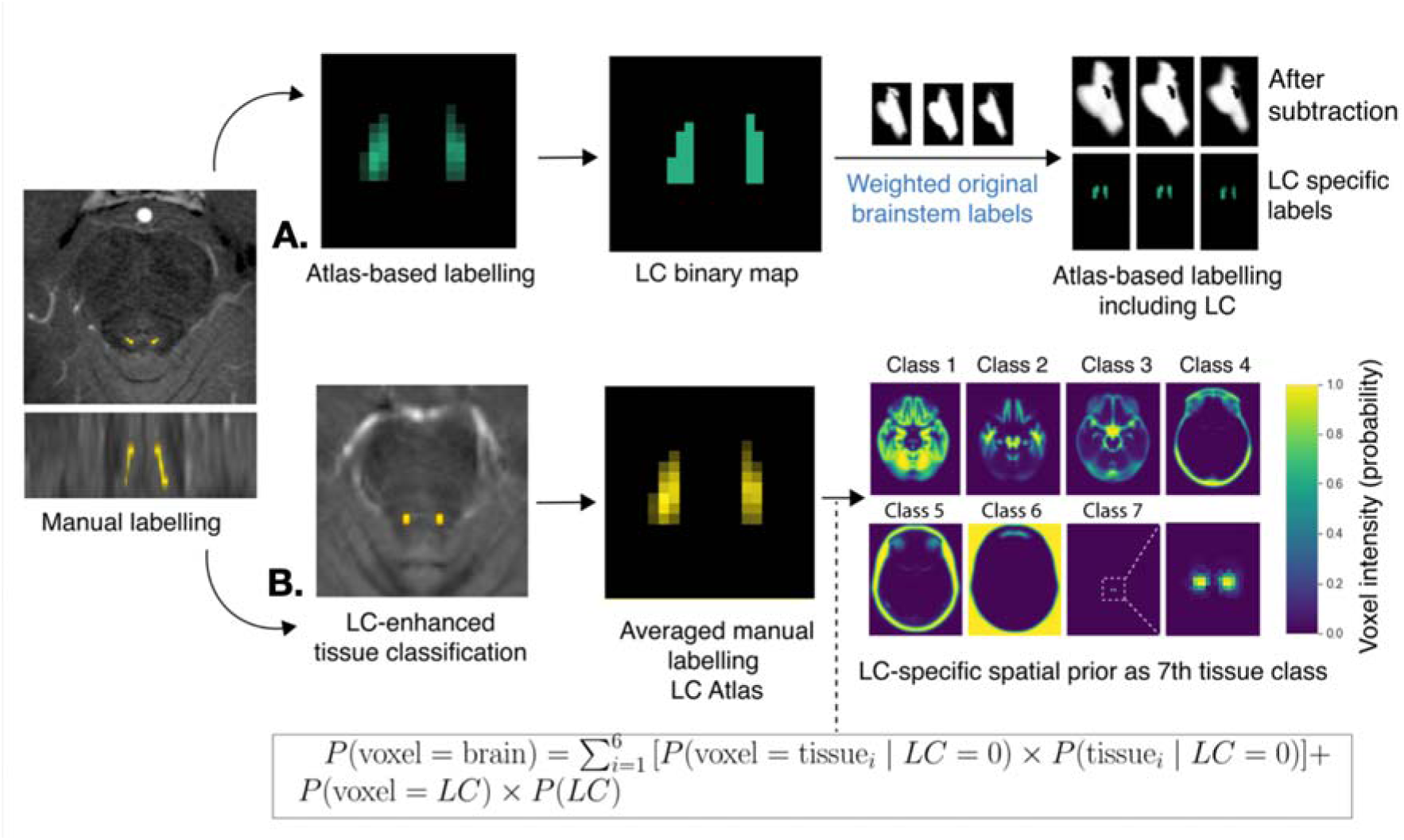
Manual LC labels from NM-MRIs used for: **A.** atlas-based labelling and **B.** SPM12s unified segmentation with LC-spatial prior. *Abbreviations: LC = locus coeruleus; NM-MRI = neuromelanin-sensitive MRI; SPM = Statistical Parametric Mapping; TPM = tissue probability map*.

Aiming to mitigate manual labelling variability, we created a consensus LC reference by combining information from the seven open-access atlases and with the averaged manual labelling LC atlas within the atlas-based labelling. We created overlap maps by determining, for each voxel, in how many atlas volumes it was present. Voxels present in two or more atlases were retained to define a consensus representation of the LC (Suppl. Fig. 2). This resulting consensus-based reference atlas was added as an additional atlas-based ROI, generating a harmonized benchmark output.

### 2.7 LC-enhanced tissue classification

We used SPM12’s unified segmentation for automated tissue classification of the NM-MRIs, incorporating a LC-specific spatial prior as an extra tissue class and NM-MRI signal intensity modelled with two Gaussians (see Suppl. Table 1A) in 1×1×1mm resolution. To assess dependence on the prior, we repeated the procedure in three ways: with each of the seven already published atlases, with the averaged manual labelling atlas or with the consensus reference atlas described above.

### 2.8 Validation

We evaluated the following features for both the spatial priors-based and the LC atlas-based labelling: (i) spatial overlap, (ii) anatomical alignment, (iii) intensity distributions and spatial patterns, and (iv) boundary distances. Spatial overlap was measured using a: (i) soft Dice, calculated by taking twice the voxel-wise product of the two probabilistic outputs, summing over all voxels, and dividing by the sum of the squared voxel values of each output, providing a continuous generalisation of the standard Dice for non-binary labels; (ii) Volume intersection, which is the ratio of intersecting voxels to the union of voxels in two outputs, capturing agreement in overall volume and (iii) voxel-wise Pearson correlation, measuring the correlation between corresponding voxel values. Anatomical alignment was assessed using the Euclidean distance between the centres of mass (COM). Intensity distributions and spatial patterns were compared using the Kullback–Leibler (KL) divergence and Earth Mover’s Distance (EMD), which determine how much and how far intensity values must be shifted to transform one spatial pattern into another without simplifying the data into binary masks. Boundary distances and outliers were quantified through the Hausdorff Distance.

We assessed the performance of the automated LC delineation approaches by measuring their spatial overlap with the corresponding consensus reference. This comparison was done in both native and MNI space, focusing specifically on the rostro-caudal dimension.

We analysed the anatomical distribution of the LC-spatial prior-based and atlas-based outputs by comparing the rostro-caudal (z-axis) and in-plane (x- and y- axis) spatial extent, considering the impact of the necessary arbitrary thresholding of the probabilistic LC maps. We used principal component analysis (PCA) on the voxel coordinates to attribute the density distributions of voxel signal along each spatial component, which we interpreted as corresponding to the anterior–posterior (PC1), left–right (PC2), and inferior–superior (PC3) axes of LC. We compared spatial dispersion across methods and used group-level statistics to test for significant differences. Analyses were conducted using NumPy and pandas for data manipulation, with visualization performed via seaborn and matplotlib and statistical testing supported by *SciPy* and *statsmodels*.

## 3 Results

### 3.1 Manual labelling

The average DSC for the manual labelling across nine raters was 0.69 ± 0.18 (mean ± SD); 95% CI: 0.67–0.72, reflecting moderate intra-rater spatial agreement. The inter-rater agreement was also moderate, with a maximum Fleiss kappa value of 0.5 (**Fig. 3**). The averaged manual labelling LC atlas (**Fig. 4)** extends rostro-caudally to 12mm in MNI space. To test the stability of the atlas creation, we performed random sub-sampling validation by creating atlases from different subsets of 21 individuals sand excluding randomly 3 individuals. The resulting atlases showed near-perfect overlap (>99%), indicating that the group atlas is highly robust to the specific subset of participants used for its creation.

**Figure 3.**
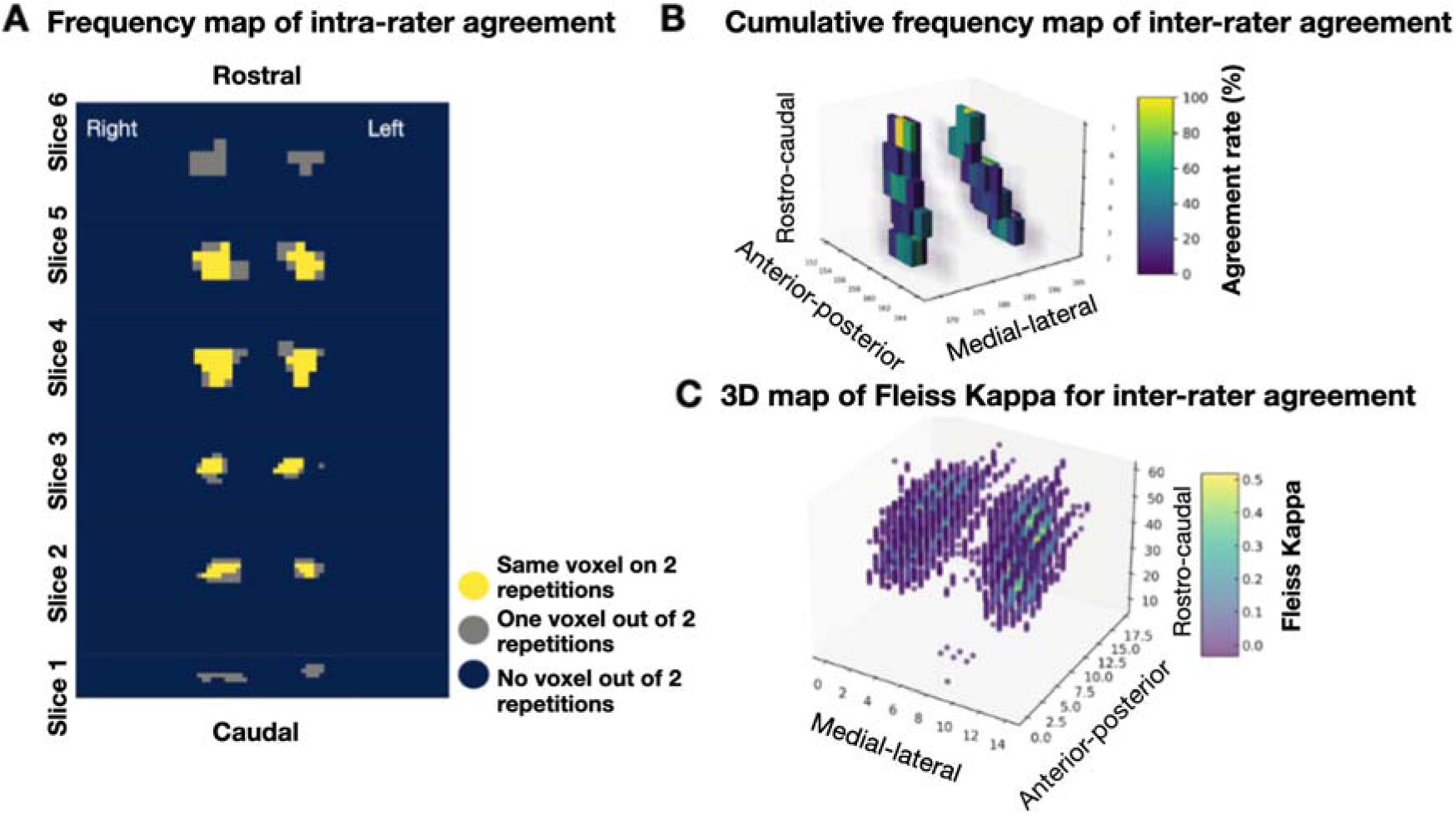
**A.** Example of intra-rater agreement maps across six rostro-caudal slices by one rater: yellow = consistent voxels, grey = single repetition, dark blue = no overlap. **B.** Cumulative frequency map of inter-rater agreement (0–100%). Colour intensity indicates relative frequency values. **C.** 3D Fleiss’ Kappa map (0–0.5) in MNI space of inter-rater reliability along rostro–caudal, anterio–posterior, and medio–lateral axes across all raters and subjects. *Abbreviation: MNI = Montreal Neurological Institute*.

**Figure 4.**
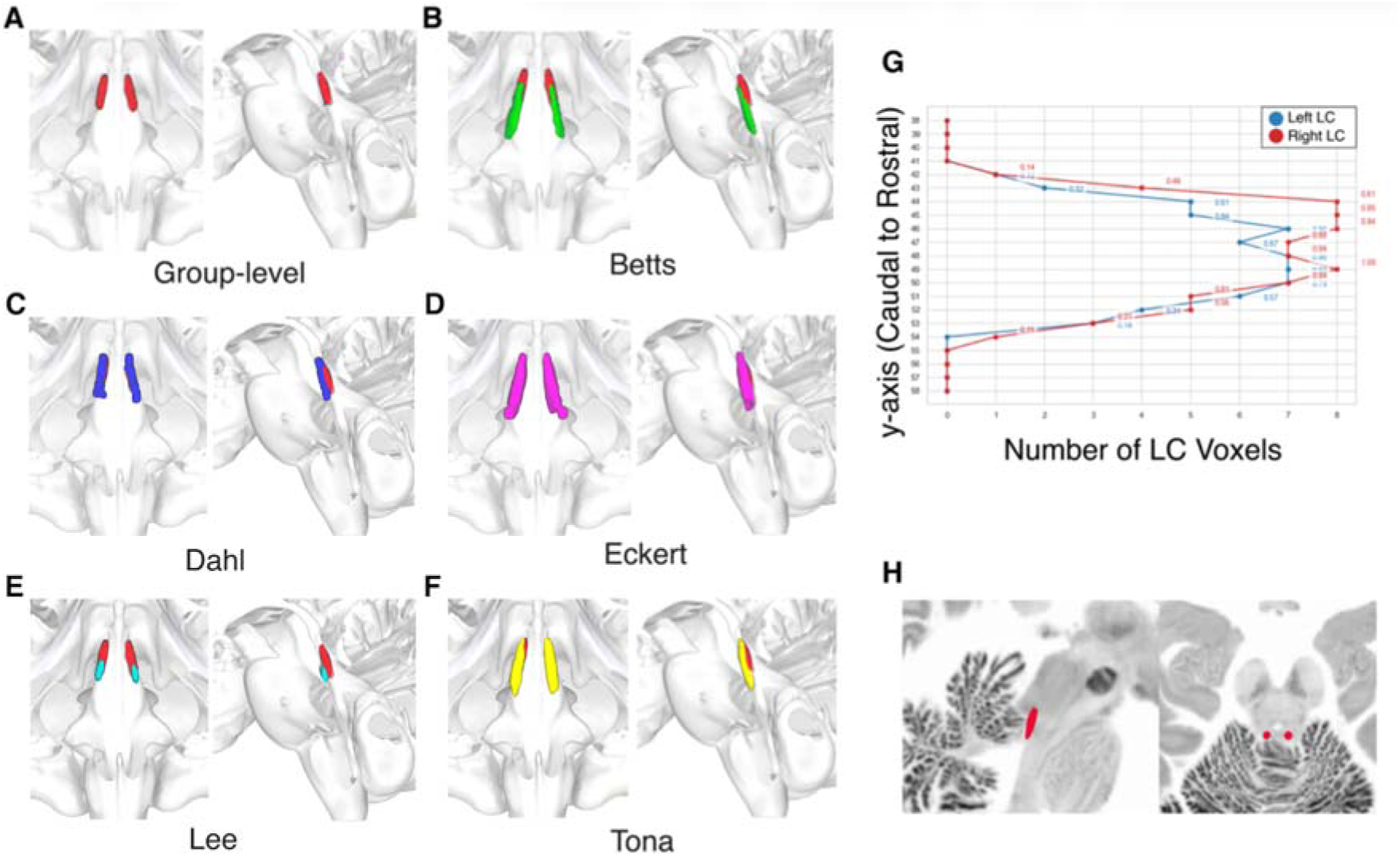
**A-F**. Renderings of LC atlases (group-level, Betts, Dahl, Eckert, Lee, and Tona) in MNI space. **G.** Number of voxels in the left (blue) and right (red) group-level LC atlas along the rostro-caudal axis. **H.** Group-level atlas projected on the BigBrain (Amunts et al., 2013). *Abbreviations: group-level = averaged manual labelling atlas; LC = locus coeruleus; MNI = Montreal Neurological Institute*.

### 3.2 Atlas-based labelling

We integrated the LC atlases into the MICCAI multi-atlas framework. To evaluate the accuracy of the atlas-based labelling, we calculated the DSC between the individual atlas and manual label for each subject. The labelling accuracy varied across authors (**Fig. 5**). DSCs ranged from low values for the Lee atlas (0.22 ± 0.07, 95% CI 0.03) to the highest for the Ye atlas (0.38 ± 0.11, 95% CI 0.04), which showed a few outliers likely due to registration errors or anatomical variability. The Tona atlas exhibited high overlap (0.25 ± 0.09, 95% CI 0.03), while the averaged manual labelling atlas had lower overlap (group-level Dice = 0.28 ± 0.09, 95% CI 0.04). Cohen’s Kappa values demonstrated a similar pattern across atlases and individuals. Volume intersection was highest for the Tona and Keren atlases (0.83 ± 0.10, 95% CI 0.03), and lowest for Lee (0.41 ± 0.09, 95% CI 0.04) and the group-level atlas (0.51 ± 0.11, 95% CI 0.04). Boundary distance measures (COM and Hausdorff) indicated that the Keren atlas produced the most consistent outputs, with the smallest distances to manual labels (Suppl. Table 2).

**Figure 5.**
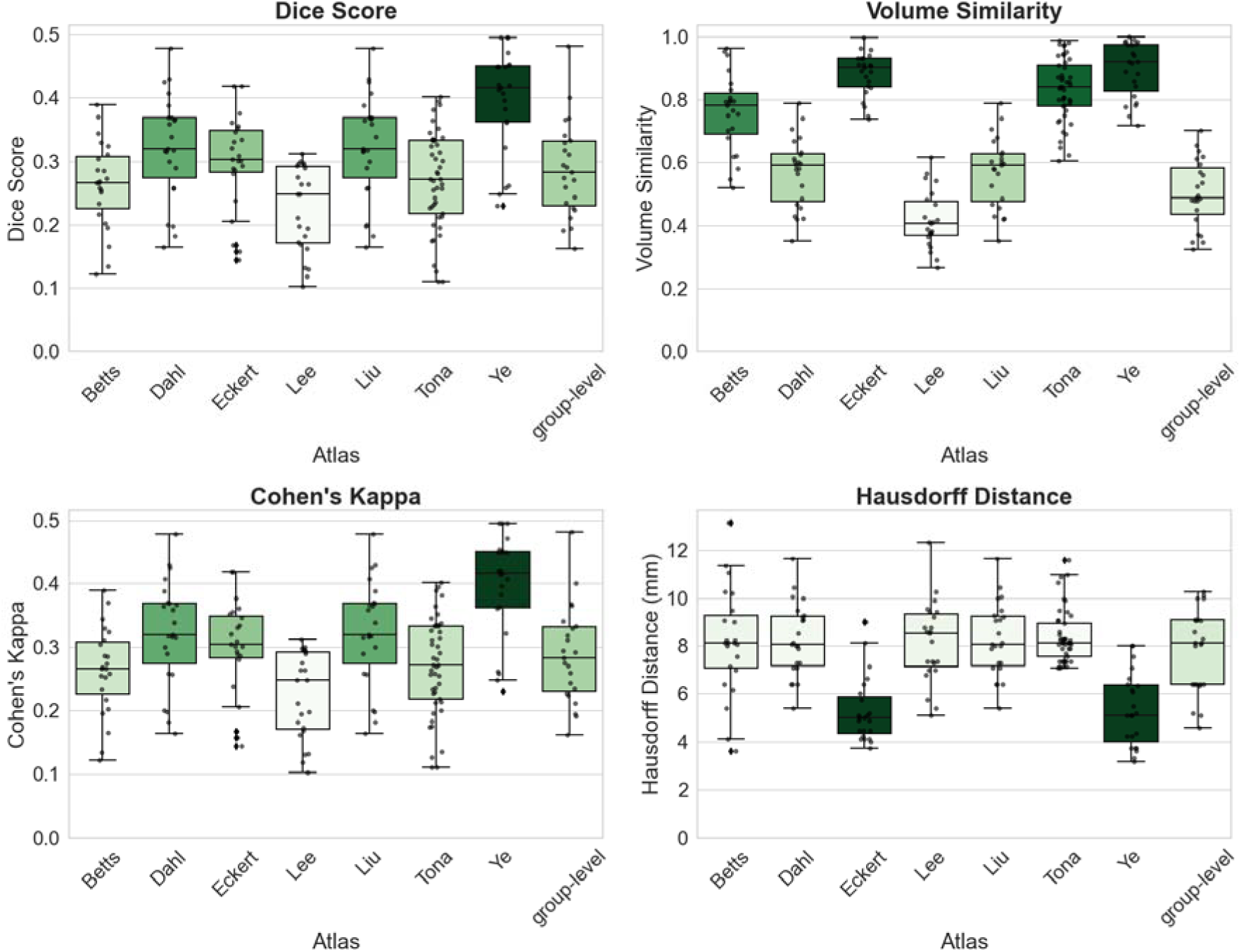
Performance comparison of atlas labelling against manual reference. Boxplots depict medians, interquartile ranges, and outliers. Darker shades indicate superior agreement with the reference manual label across all metrics. For the Hausdorff Distance, lower values correspond to darker shades. *Abbreviations: group-level = averaged manual labelling atlas*.

The analysis comparing atlas-based labels to the consensus reference (Suppl. Fig. 3) showed that the averaged manual labelling atlas had the highest spatial concordance (mean Dice coefficient = 0.63 ± 0.02, 95% CI 0.01), while the Tona and Liu atlases had the lowest. Volume similarity was generally high across most atlases, with the Lee atlas at the lowest performance range. Labels derived from the Ye and Keren atlases achieved the closest boundary distances to the consensus reference atlas, whereas the Tona atlas showed the greatest differences.

### 3.3 LC-specific spatial prior based delineation

In SPM12’s tissue classification with a LC-specific prior using group-level and Dahl atlas as showed the most accurate and spatially consistent performance relative to manual labels in native space (**Fig. 6**). Using the Dahl atlas as a prior, we obtained the highest mean Soft Dice coefficient (0.46 ± 0.14, 95% CI 0.06) and volume intersection with the manual labels (90.57 ± 31.31, 95% CI 12.80). In contrast, using the group-level atlas as LC prior achieved the highest voxel-wise correlation (0.33 ± 0.21, 95% CI 0.08) and the lowest mean KL divergence (3.91 ± 2.47, 95% CI 1.01), indicating better overall spatial consistency. The COM distance analyses showed outliers in four out of the eight priors, with the Betts atlas-based prior showing the greatest variability (6.47 ± 8.58, 95% CI 3.50). Evaluation of KL divergence showed the group-level LC prior achieved the lowest mean divergence (3.91 ± 2.47, 95% CI 1.01). EMD was lowest for Dahl- and Ye atlas based priors (Dahl atlas: 3.88E-08 ± 1.88E-08, 95% CI 7.67E-09; Ye: 6.77E-08 ± 2.86E-08, 95% CI 1.17E-08), though the Ye atlas based prior showed greater variability across subjects (for details see Suppl. Table 3).

**Figure 6.**
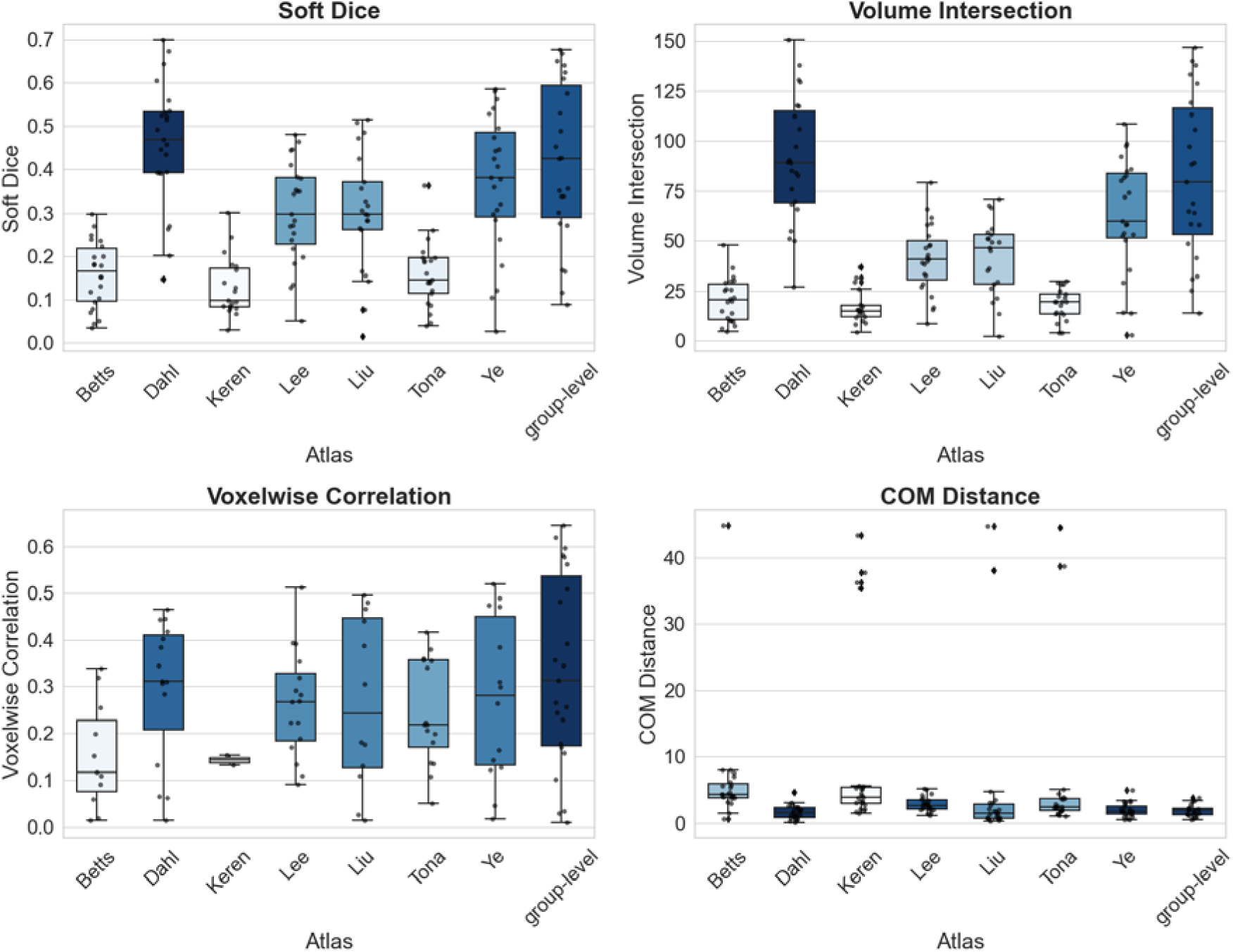
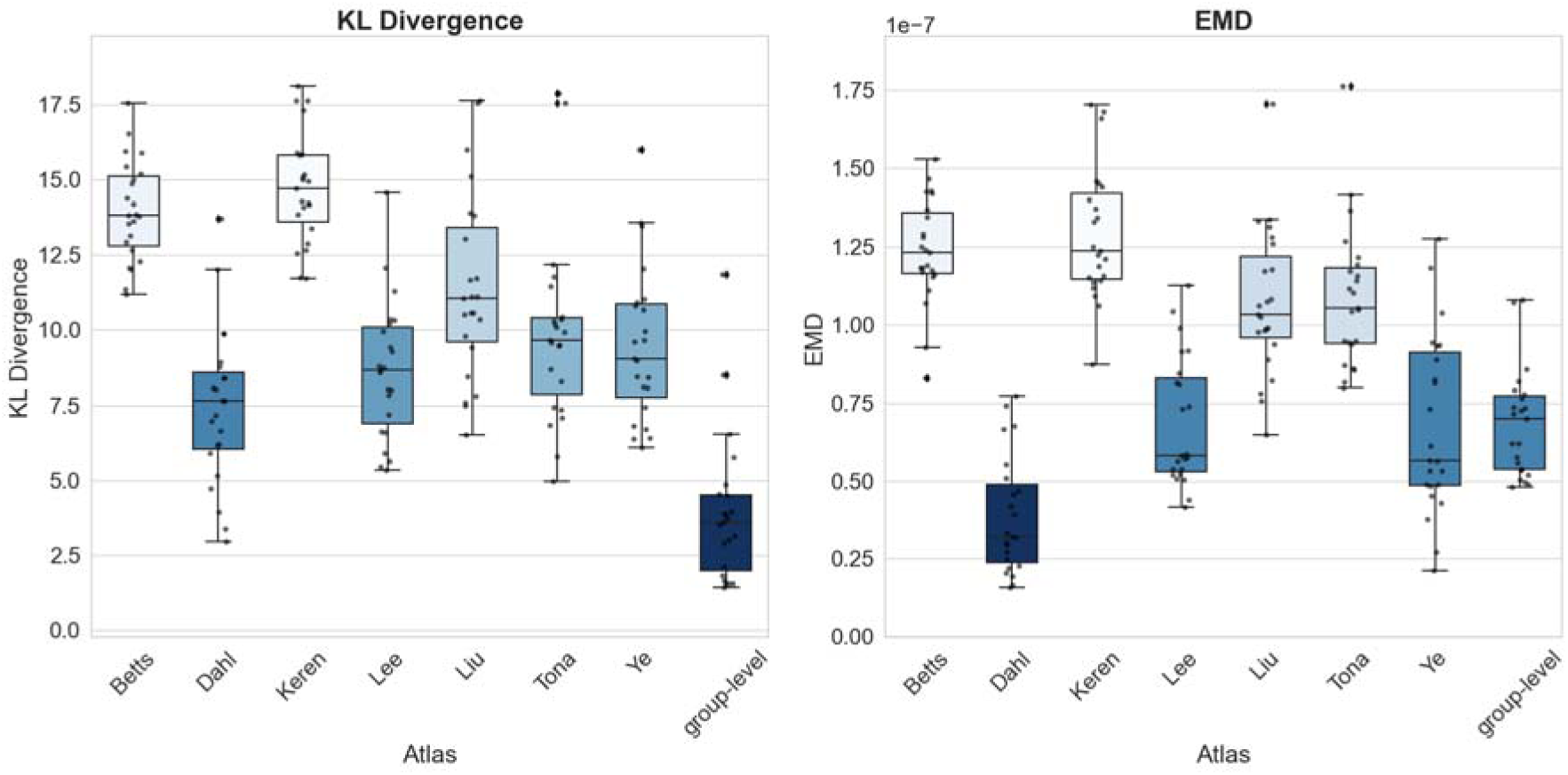
Comparison of LC-specific spatial prior-based delineation against manual labels in native space. Boxplots display medians, interquartile ranges, outliers and individual data points. For all metrics, colour intensity reflects performance, with darker shades indicating greater agreement with the reference manual label. *Abbreviations: COM = centre of mass; EMD = Earth mover’s distance; group-level = averaged manual labelling atlas; KL divergence = Kullback–Leibler divergence*.

We compared the created LC maps from the automated tissue classification against the consensus reference (Suppl. Fig. 4). The performance trends across authors observed for the Soft Dice and Volume Intersection remained consistent with those obtained from comparisons to manual labelling. Voxel-based correlation values decreased for all outputs in this comparison, except when using the group-level atlas as LC prior, which maintained higher correlation levels. The COM distance demonstrated a broader distribution of values compared to manual labelling assessments.

### 3.4 Spatial distribution and concordance of LC delineation methods

To compare the consistency of LC delineation methods, we evaluated the spatially aligned outputs of atlas-based labelling and LC-specific spatial prior delineation in terms of cross-sectional area along the rostro–caudal axis (**Fig. 7**). Both methods localized the structure within a comparable rostro–caudal extent, though the atlas-based labelling generally exhibited greater cross-sectional area across slices and hemispheres. The third column summarizes the eight LC atlases, depicting the mean cross-sectional area (lines) and ±1 SD variability (shaded) as a visual reference for the atlas inputs and their transformation in the delineation outputs.

**Figure 7.**
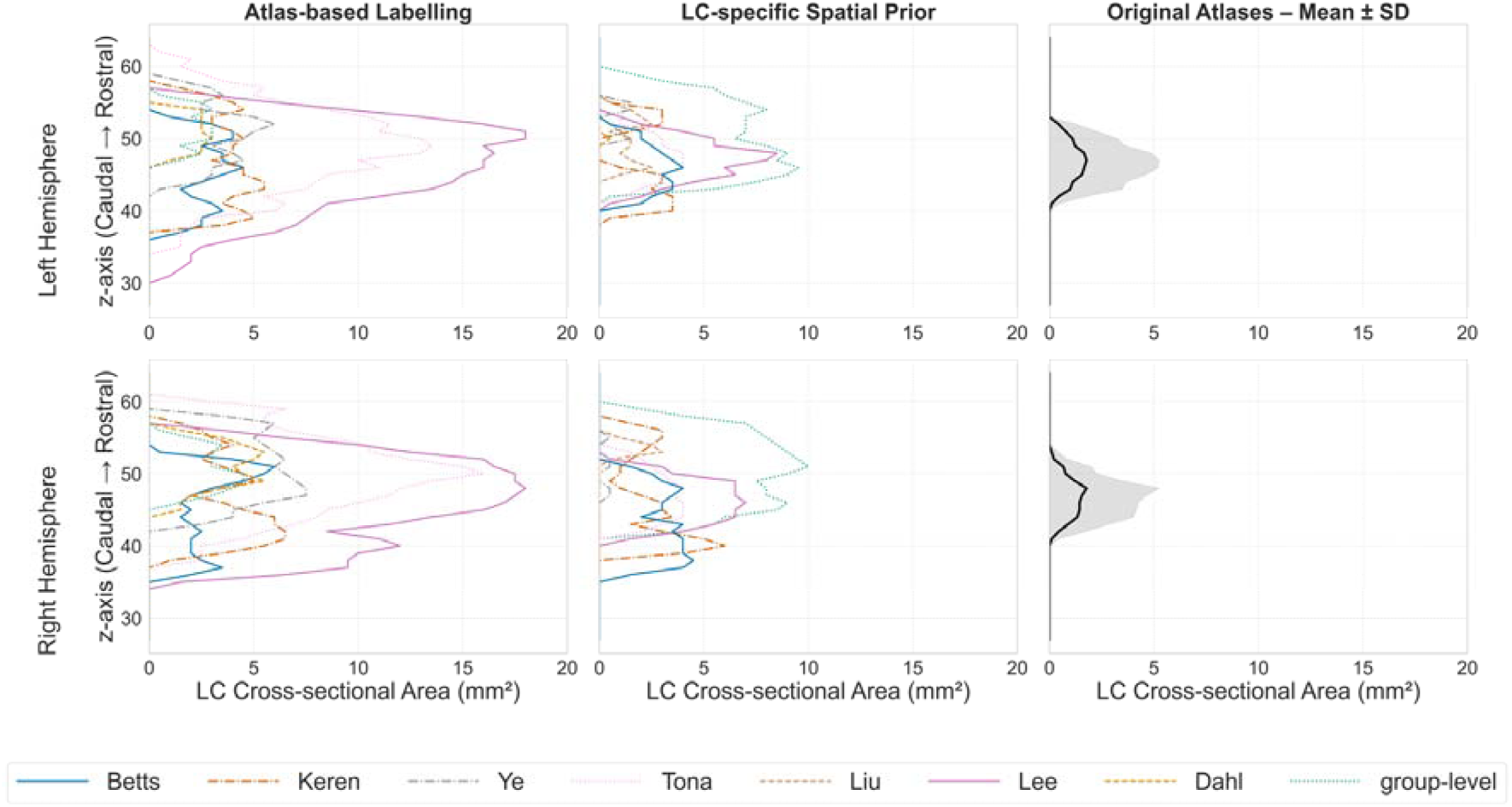
Cross-sectional LC area along the rostro–caudal (z-) axis in MNI space. The first two columns display per-author results for the atlas-based labelling and LC-focused spatial tissue prior methods, where each line represents an individual author mask. The third column summarizes the original LC atlases, showing the mean cross-sectional area (black line) and ±1 SD variability (grey shading). *Abbreviations: group-level averaged manual labelling atlas; LC = locus coeruleus; SD = Standard Deviation*.

Given the variation in spatial overlap between delineation approaches, particularly along the rostro–caudal LC axis, we next evaluated their reliability in localizing the LC. The atlas-based labelling showed generally higher agreement in standard space, particularly for the Lee, Tona and Ye atlases, which achieved near-perfect overlap (>95%) across all three rostro–caudal LC tiers (see Suppl. Figs. 5 and 6). On the contrary, the overlap was consistently lower and more variable across atlases in native space, with especially low agreement in the rostral section (Tier 1) when dividing the LC in three tiers (Suppl. Fig. 7). Collapsing the LC to 2 tiers slightly improved average overlaps but did not resolve the spatial inconsistencies (Suppl. Fig. 8).

The LC delineation based on spatial priors showed lower overlap with the consensus reference (Suppl. Figs. 9-12), particularly in native space, where alignment across tiers was poor. Only the group-level and the Lee-based LC prior demonstrated high spatial overlap in MNI space, with the group-level atlas approximating 90% in Tier 2. Reducing the search volume from three to two tiers improved consistency in some cases but did not substantially elevate overall agreement.

To summarise the spatial organization of detected LC voxels across the two delineation approaches, we applied a PCA to the outputs from atlas-based labelling and LC-enhanced tissue classification in MNI space. PCA revealed three major axes of spatial variance, approximately aligned with the anatomical left–right (PC1), caudal–rostral (PC2) and anterior–posterior (PC3) directions. The PCA component coordinates in MNI space were: PC1 = [0.914, 0.396, 0.091], PC2 = [0.098, -0.431, 0.897], PC3 = [-0.395, 0.811, 0.433]. The angular deviation between the PCA and AC–PC coordinate systems was [23.99°, 64.45°, 64.36°] for PC1, PC2, and PC3. The orientation of the PCA axes suggests that the LC is elongated along an oblique axis relative to the canonical AC–PC frame, in line with its anatomical rotation in the brainstem. Density plots for each principal component (**Fig. 8, top row**) showed spatial dispersion differences between approaches. The LC-specific spatial prior-based delineation showed broader voxel distributions across all components, particularly along the left–right (PC1) and caudal–rostral (PC2) axes, indicating a more spatially diffuse representation. In contrast, outputs of the atlas-based labelling were narrowly distributed and centred around zero, suggesting greater spatial coherence. The statistical testing confirmed significant differences between approaches along PC1 (p = 2.00 × 10^−3^, **), PC2 (p = 4.00 × 10^−2^, *), but not PC3 corresponding to the caudal–rostral direction (p = 1.50 × 10^−1^, ns). The voxel coordinate projections in PCA space (**Fig. 8, bottom row**) showed the differences in spatial delineation accuracy. The scatter plots confirm these patterns: atlas-based labelling clustered tightly in all planes, whereas spatial prior–based delineation showed greater dispersion, particularly along PC1 and PC2, indicating higher spatial variability.

**Figure 8.**
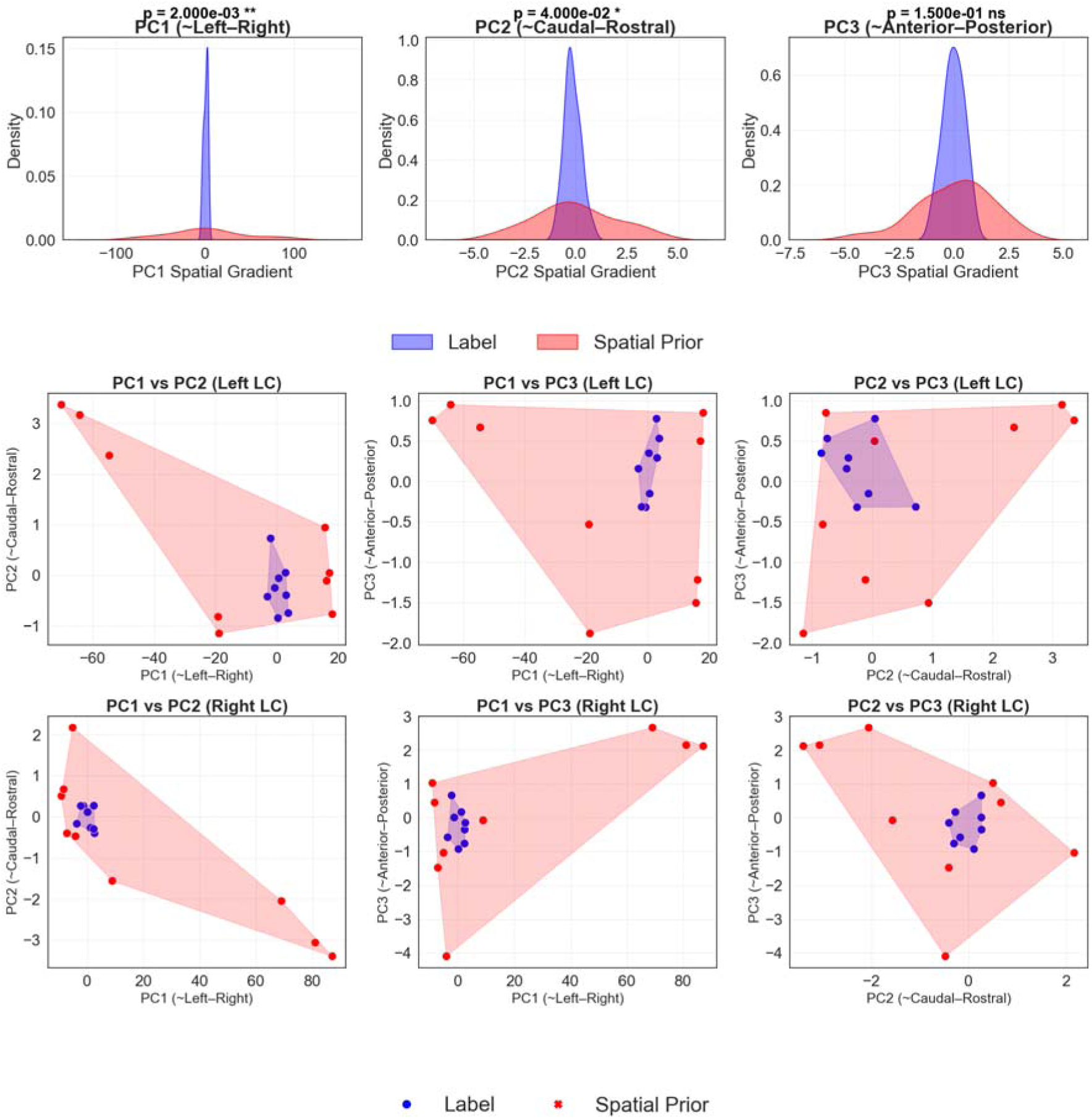
Voxel-wise spatial gradients across principal components (PCs) for atlas-based labelling (blue) and LC-spatial prior based tissue classification (red) in MNI space. Kernel density estimates for PC1 (left–right), PC2 (rostro-caudal), and PC3 (anterio–posterior). Scatter plots of paired PCs for left and right LC. *Abbreviations: group-level = averaged manual labelling atlas; PC = Principal Component; ns = non-significant; * = <0.05, ** = <0.002*.

## 4 Discussion

In this study, we compared atlas-based, and spatial prior-guided LC delineation to manual labeling of NM-MRI data to examine how methodological choices influence spatial accuracy and consistency in both native and standard reference spaces.

Atlas-based labelling, when using the group-level manual labels atlas, produced the most accurate and spatially coherent LC representations in both native and standard space. This atlas consistently achieved the highest concordance with the manual labels and the consensus reference to show lower spatial variability than LC-specific spatial prior-based delineation. The Ye and Keren atlases also performed well in specific metrics, such as boundary distance, but no single atlas excelled across all measures. These results highlight the value of consensus-based and group-level atlases for achieving robust LC segmentation, especially when aiming for anatomical consistency across studies.

The obtained method-depending differential LC delineations are consistent with previous research reporting that structural and functional LC localization approaches identify overlapping but not identical LC regions (Mäki-Marttunen & Espeseth, 2021). Atlas-based labelling produced more spatially coherent LC maps, with principal component analysis indicating tighter clustering and clearer separation along anatomical axes compared to the more diffuse spatial prior-based method, particularly along the left–right and rostro-caudal axes. The broader volume extent observed in the spatial prior–based approach likely reflects thresholding uncertainty inherent to probabilistic maps, which penalises tissue classification when evaluated with binary metrics (Trutti et al., 2021). Nonetheless, the lack of statistically significant differences along the anterio–posterior LC axis suggests partial anatomical convergence between approaches. Thus, atlas-based labelling may be better suited for studies prioritising spatial precision, while tissue classification could offer advantages in applications involving uncertainty modelling.

Despite generally high overlap metrics across individual atlases, boundary variability was common across metrics. These inconsistencies likely stem from differences in atlas creation methods and demographic characteristics. These methodological differences shaped the nature of the atlas maps: manually labelled atlases produced deterministic maps with fixed boundaries, while those derived from semi-automated, intensity-based methods produced smoother probabilistic maps. Differences in study populations of the included atlases further contributed to variability. Given that neuromelanin accumulates with age, LC contrast may be lower in younger individuals, limiting delineation sensitivity and potentially biasing atlas construction (Berger et al., 2023; Ohm et al., 1997). Most atlases used contrast or signal-to-noise ratio for LC identification, but only two accounted for age-related effects, and one used lower-resolution imaging. To ensure comparability, all atlases were down sampled, which limits the anatomical precision. These factors should be considered when interpreting differences across atlases. However, formal comparison of how these factors impacted each atlas was beyond the scope of this study.

In such small volumes as the LC, even minor boundary mismatches substantially reduce Dice scores. Consensus references, akin to majority-vote approaches (Ye et al., 2021), can minimise individual labelling bias and strengthen cross-atlas comparisons. Aggregating input from multiple raters, even when individual maps are sparse and variable, can enhance robustness by smoothing out idiosyncratic differences and emphasising consistently labelled regions. The result is a more generalisable and anatomically coherent LC representation that better captures common structural features across individuals.

Following registration into MNI space, improvements in spatial correspondence were most evident in the central portion of the LC. This finding aligns with known LC properties in healthy individuals, where the central LC often exhibits stronger signal and less inter-individual variability. Prior studies have also shown that LC-related signal alterations, due to aging, depression, or early Alzheimer’s disease, are often most prominent in the central or rostral-middle portions (Bae et al., 2025; Hary et al., 2025; Shibata et al., 2007). These physiological and pathological variations help explain some atlas discrepancies. Differences in atlas coverage have been analysed, with the Dahl atlas underrepresenting the caudal LC and the Betts atlas being less extensive rostrally (Liu et al., 2019). Taken together, these spatial differences highlight the importance of considering intra-LC variability when interpreting atlas performance and LC delineation results.

The observed spatial variability in native space reinforces the importance of using optimized registration algorithms for small nuclei. The high variability and number of outliers, especially in rostral and caudal parts of the LC, highlight the limits of manual labelling and the need for harmonised references. This aligns with previous work supporting group-level LC atlases, tailored to the characteristics of the study cohort, showing greater localisation precision compared to atlases derived from independent populations (Mäki-Marttunen & Espeseth, 2021). Altogether, these findings emphasise the value of adopting context-sensitive approaches.

Several limitations should be considered when interpreting findings of this study. Although manual labelling is often treated as ground truth in LC imaging (Doppler et al., 2021; Mäki-Marttunen & Espeseth, 2021) or as correction after automated segmentation (Wolters et al., 2023), our results highlight its inherent variability and limited spatial consistency with Dice scores based on comparisons between manual and automated methods––below 0.4, particularly at the rostral and caudal ends. Inter-operator discrepancies (Daboul et al., 2018) and anatomical ambiguity aggravated by low contrast in neuromelanin-sensitive MRI may have introduced noise into the manual reference labelling, potentially affecting all comparative evaluations. The small sample size (n = 24) in this study may limit the generalisability of the results and the robustness of some comparisons. Moreover, the representativeness of the sample in terms of socio-demographic characteristics was not systematically assessed, and the robustness of findings across different scanners or patient groups with varying disease profiles remains to be established.

Our study achieved a mean inter-rater reliability of 0.7, aligning with previously reported values in the literature (54–80%) (Betts et al., 2019; Liu et al., 2017; Tona et al., 2017). Second, while we used down-sampled versions of available LC atlases to maintain consistency, this may have reduced the granularity of finer LC features present in the original high-resolution templates. Third, the consensus reference atlas required voxel-wise binarization, which assumes equal atlas quality and may underrepresent high-confidence voxels from individual atlases. This approach, though practical, assumes equal quality and anatomical accuracy across all input atlases and may underrepresent LC voxels with high confidence from single high-quality sources. Lastly, we did not evaluate deep learning-based approaches, which are increasingly used in LC delineation. Although outside the scope of this study, such methods offer a promising direction for future work, particularly in large-scale or clinical applications.

We conclude that LC identification in structural MRI remains a complex task, requiring a careful balance between delineation approach, atlas selection and validation strategy. Manual labelling remains a common reference in smaller cohorts, yet the inherent variability and limited reproducibility challenge its role as a definitive ground truth. Automated and atlas-based approaches offer significantly greater scalability when benchmarked against a consensus reference atlas. Our study highlights the relative advantages of atlas-based labelling for achieving spatial precision independent of MRI contrast, allowing for the development of reproducible LC imaging biomarkers in future studies. Probabilistic LC priors provide flexibility in representing anatomical uncertainty when neuromelanin-sensitive imaging is used. Among the eight atlases evaluated, the manually based group-level LC atlas demonstrated robust and generalisable performance across selected statistical metrics and delineation approaches. Our results support atlas-based labelling as a reliable and anatomically consistent approach for LC identification, particularly in combination with harmonised consensus-based reference frameworks. Finally, these findings emphasise the importance of method-appropriate validation metrics and the ongoing need for standardisation in neuromelanin-sensitive imaging.

## 5 Data and Code Availability

The data from the CoLaus|PsyCoLaus cohort used in this study cannot be fully shared due to the inclusion of potentially sensitive patient information. According to the competent authority, the Research Ethic Committee of the Canton of Vaud, Switzerland, sharing or transferring this data would violate Swiss legislation designed to protect participants’ personal rights. However, non-identifiable, individual-level data can be made available to researchers who meet the criteria for accessing confidential data, for detailed instructions please see https://www.colaus-psycolaus.ch/professionals/how-to-collaborate/) by CoLaus|PsyCoLaus Datacenter (CHUV, Lausanne, Switzerland). Code to all analyses and figures will be uploaded to https://github.com/dmiolga/LC_delineation.git once the paper has been accepted.

## 6 Author Contributions

**Olga Dmitrichenko**: Conceptualization, Methodology, Validation, Formal Analysis, Visualization, Investigation, Writing - Original Draft, Writing - Review & Editing, Visualization. **Giuseppina Baldizz**i: Data Curation, Methodology, Validation, Formal Analysis, Visualization, Writing - Original Draft, Writing - Review & Editing. **Tatjana Schmidt**: Data Curation, Writing - Review & Editing. **Aurélie Bussy**: Methodology, Writing - Review & Editing. **Olivier Colliot**: Methodology, Writing - Review & Editing. **Antoine Lutti**: Data Curation, Resources, Writing - Review & Editing. **Ferath Kherif**: Methodology, Resources, Writing - Review & Editing. **Bogdan Draganski**: Supervision, Conceptualization, Data curation, Funding acquisition, Investigation, Methodology, Project administration, Resources, Writing - Original Draft, Writing - Review & Editing. All authors read and approved the final manuscript.

## Acknowledgments and Funding

B.D. is supported by the Swiss National Science Foundation (project grant no. 213595, 32003B_135679, 32003B_159780, 324730_192755 and CRSK-3_190185),

ERA_NET NEURON JTC2020: iSEE and JTC2023-ELSA: BrainTree projects and the InnoSuisse Flagship Swiss brAInHealth project. A.L. is supported by the Swiss National Science Foundation (grant no. 320030_184784, CR00I5-235940). The Laboratory for Research in Neuroimaging-LREN is very grateful to the Roger De Spoelberch and Partridge Foundations for their generous financial support.

## 8 Supplementary Material

Figs. S1 to S12

Tables S1 to S3

