## Supplementary Material for "The quest for the best: manual, atlas- and spatial prior-based delineation of locus coeruleus"

**MRI data pre-processing**

Magnetic resonance imaging (MRI) data preprocessing was performed in the framework of Statistical Parametric Mapping (SPM12; <http://www.fil.ion.ucl.ac.uk/spm>; Wellcome Centre for Human Neuroimaging, University College London, UK). For tissue classification of the T1-weighted data, we used the “unified segmentation” with enhanced tissue priors (Lorio et al., 2016). Then, we performed a two-step diffeomorphic spatial registration to the MICCAI-labels space (Yan et al., 2022) using geodesic shooting (Ashburner & Friston, 2011) followed by registration to the standard Montreal Neurological Institute (MNI152) space.

The neuromelanin-sensitive MR images (NM-MRI; n=24) were co-registered to the individuals’ T1-weighted MRIs using a 6-parameter rigid-body registration with nearest neighbor interpolation. In this step, we resampled the NM-MRIs from their original voxel size (0.51×0.51×3.24 mm) to an isotropic voxel resolution of 1mm³. The MN-MRIs were then subjected to the same two-step spatial registration using the diffeomorphic registration parameters from the previous iteration.

Following the manual labelling on the NM-MRIs (n=24) performed twice by 9 independent raters, we built a group-level atlas using as input the binary manual labels resulting in average frequency maps. The resulting maps were co-registered, resampled to 1mm³ using trilinear interpolation, spatially registered using the already estimated diffeomorphic registration parameters and finally averaged across participants. Low-agreement voxels were removed using Otsu’s method followed by normalization of voxel intensities to the range [0, 1]. At each preprocessing step, image quality and spatial alignment were visually assessed using SPM12 and the software MRIcroGL (version 1.2.20220720; <https://www.nitrc.org/projects/mricrogl>).

**Tissue classification of NM-MRI data using explicit locus coeruleus spatial prior**

Aiming for an automated delineation of the locus coeruleus (LC) from NM-MRIs, we included to the SPMs six tissue priors a seventh tissue class spatially encoding the LC location probability. The seven publicly available and the manually derived group-level LC atlases used as spatial priors were first rescaled in a way that the voxel intensities were between 0 and 1, where the maximum value (0.284) was set to 1. This ensured an intensity range that corresponded to the LC area in subsequent analyses. To further enhance specificity, Otsu’s image thresholding was applied. A value of 0.27 was identified as the optimal threshold and applied to the normalized probability mask, preserving only voxels with intensities above this threshold. Given the differing voxel dimensions and deterministic rather than probabilistic nature of other available LC atlases, specific adjustments were necessary to facilitate their integration into the Unified Segmentation framework.

For each voxel with non-zero LC probability, the remaining probability of SPMs tissue priors (1 – LC probability) was proportionally redistributed among the six existing tissue classes based on their original distributions. This ensured that the total probability per voxel remained normalized to 1, preserving the internal consistency of the segmentation. The tissue priors were resampled to 1mm^3^ spatial resolution. We determined the number of signal Gaussians to 2, while keeping the other unified segmentation default parameters.

**Atlas-based LC labels**

We integrated the eight open-access LC atlas information as additional label to the "MICCAI 2012 Grand Challenge and Workshop on Multi-Atlas Labeling" (Landman and Warfield, 2012; [www.neuromorphometrics.com](http://www.neuromorphometrics.com)) framework. Given the fact that the MICCAI labels space is not aligned to MNI standard space, we estimated diffeomorphic spatial registration parameters between the MICCA and MNI spaces. To ensure consistency, we also resampled all LC atlas-based labels to a 1.5x1.5x1.5mm spatial resolution. This was followed by signal intensity thresholding using Otsu’s method and binarization.

Due to variations in the structural characteristics of the included LC atlases, such as differences in voxel resolution, probabilistic representation, and challenges associated with upsampling imposed by the compact and spatially constrained anatomy of the LC, methodological adaptations were required to enable the integration of these atlases into the pipeline. The atlas-based parcellation was then performed on T1-weighted images.

**Principal Component Analysis of LC Voxels**

To quantify the spatial organization of LC voxels across the two delineation approaches, we extracted voxel coordinates from consensus maps generated by atlas-based labelling (“label”) and LC-enhanced tissue classification (“spatial prior”). Only voxels exceeding the 0.01 threshold were considered. For each approach, coordinates were transformed into MNI space and aggregated across subjects.

A principal component analysis (PCA) was performed on the stacked voxel coordinates for each approach using the *scikit-learn* implementation, retaining the first three principal components. These components captured the primary axes of spatial variance across voxels. PCA component vectors and angular deviations relative to the standard AC–PC axes were recorded to assess alignment with anatomical directions. The principal components were approximately aligned as follows:

- PC1 (~Left–Right)
- PC2 (~Caudal–Rostral / Inferior–Superior)
- PC3 (~Anterior–Posterior / Depth)

|  | PC1 | PC2 | PC3 |
| --- | --- | --- | --- |
| x | 0.914 | 0.098 | -0.395 |
| y | 0.396 | -0.431 | 0.811 |
| z | 0.091 | 0.897 | 0.433 |

The mean PCA projections for each subject and approach were extracted for downstream analysis and visualization.

**Visualization**

We visualized the PCA results using multiple complementary approaches:

1. **Density Plots**: Kernel density estimates (KDE) were computed along each principal component, comparing Label and Spatial Prior distributions. Statistical differences between approaches were annotated with p-values derived from two-sample t-test.
2. **Scatter Plots with Convex Hulls**: Pairwise projections of PCA components (PC1 vs. PC2, PC1 vs. PC3, PC2 vs. PC3) were plotted for the left and right locus coeruleus. For each approach convex hulls highlighted the full spatial extent of voxel distributions.

All visualizations were generated using Python packages *numpy, pandas, nibabel, scikit-learn, matplotlib, and seaborn*, with consistent color schemes (blue for Label, red for Spatial Prior). This PCA analysis confirmed that atlas-based labelling produces spatially tighter and more coherent LC maps, whereas the spatial prior approach exhibits broader, more diffuse voxel distributions along left–right and caudal–rostral axes, consistent with probabilistic map variability.

### **Figures**

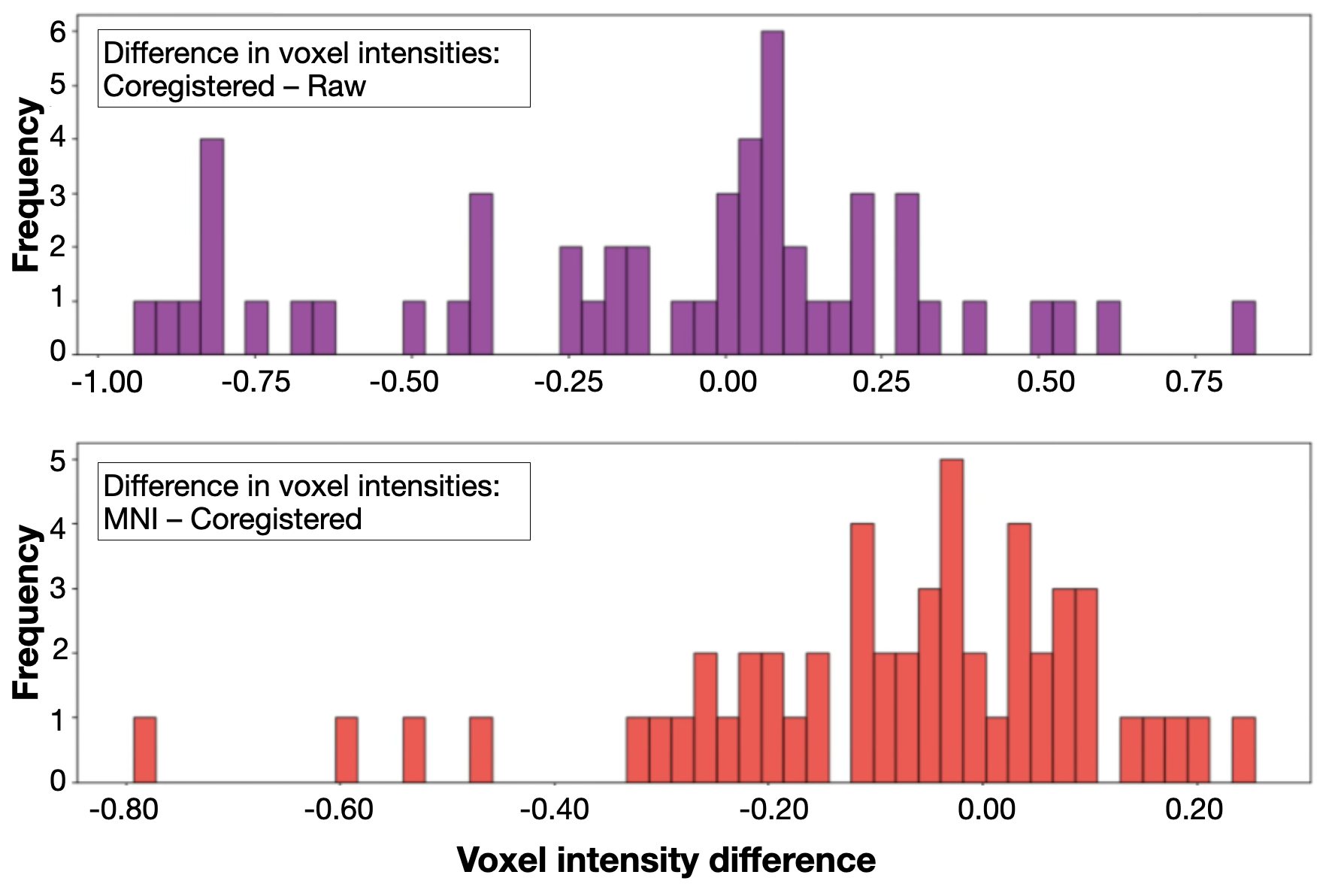
**Supplemental Figure 1**. **Histograms of voxel intensity differences across registration steps.** *Top panel*: Voxel intensity differences between coregistered and raw images. *Bottom panel:* Voxel intensity differences between spatially transformed images to MNI and their corresponding coregistered versions.

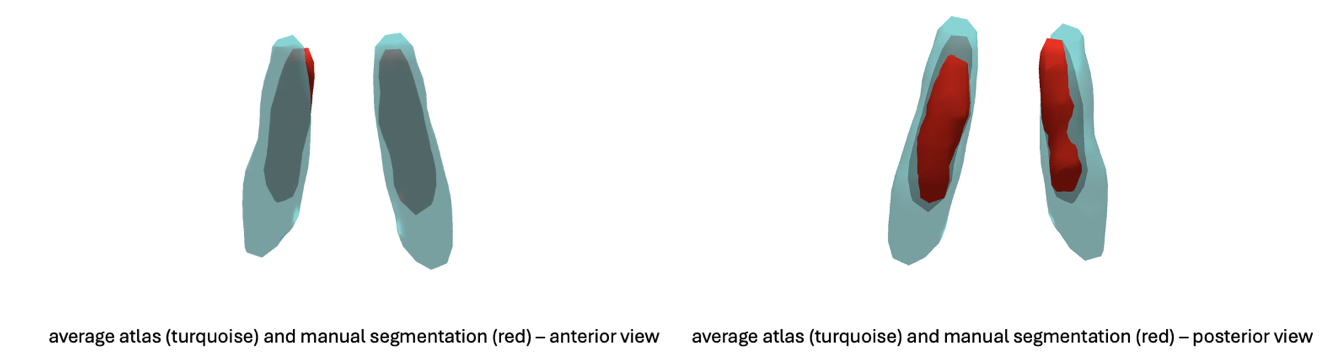

**Supplemental Figure 2**. Manually derived group level LC atlas (red) overlaid on the consensus reference atlas (turquoise)

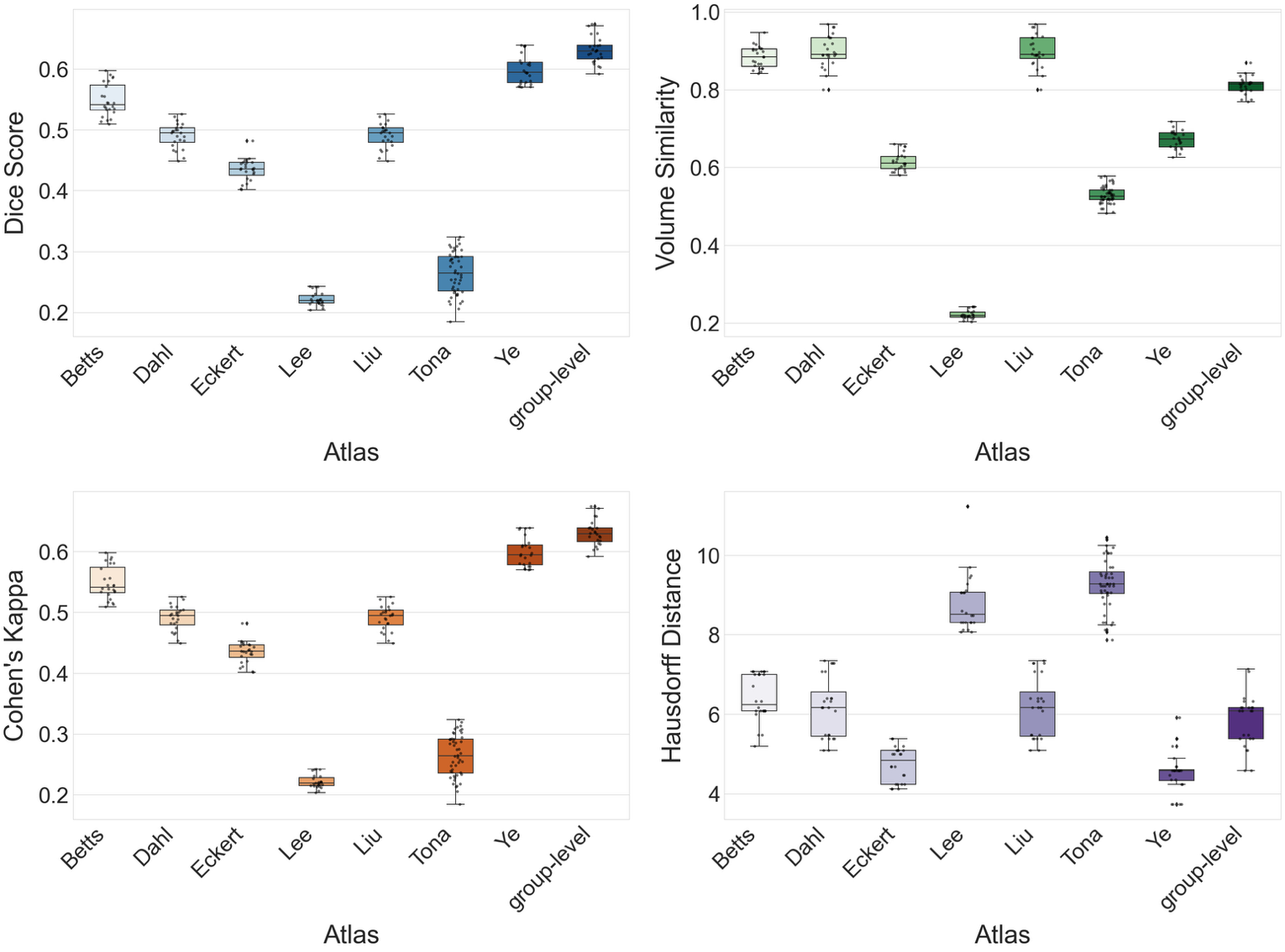

**Supplemental Figure 3.** Atlas-based labelling output in native space compared with the consensus reference. Boxplots depict medians, interquartile ranges, and outliers.

*Abbreviations: group-level = manually derived group-level atlas.*

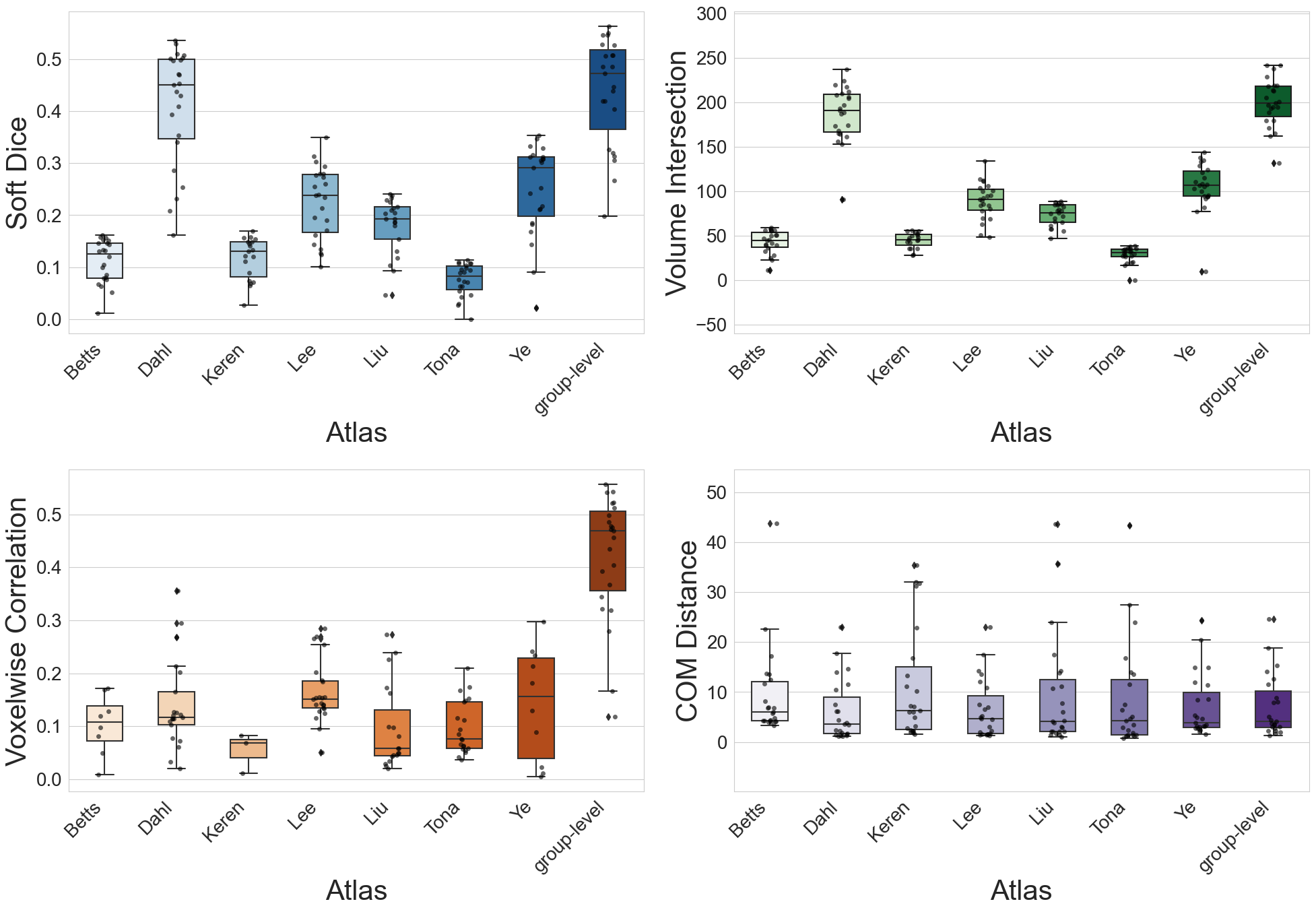

**Supplemental Figure 4**. LC-enhanced tissue classification output in native space compared with the consensus reference. Boxplots depict medians, interquartile ranges, and outliers.

*Abbreviations: group-level = manually derived group-level atlas.*

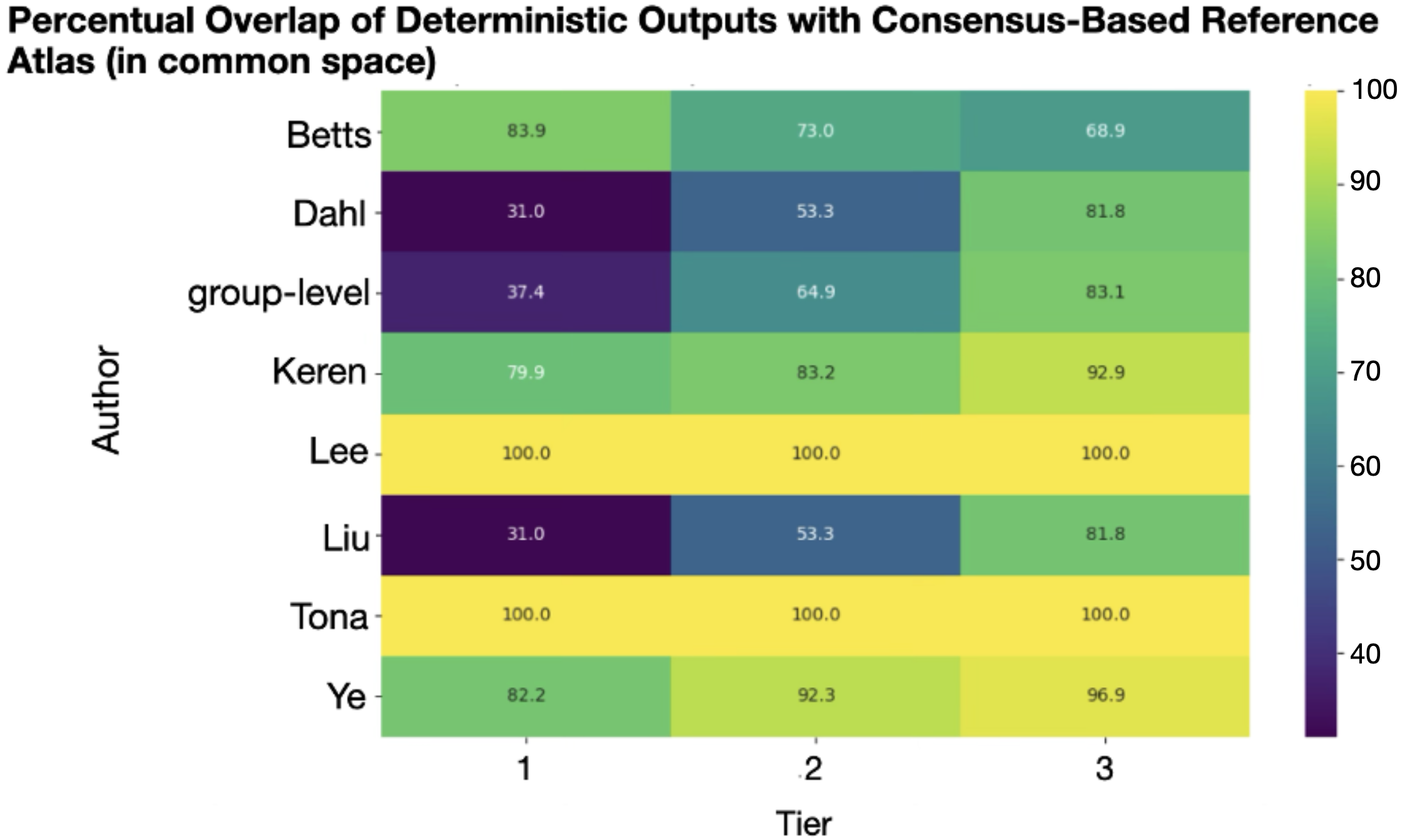

**Supplemental Figure 5**. Overlap (in %) of atlas-based parcellation with the consensus reference atlas in MNI space (3 tier division of the LC).

*Abbreviations: group-level = manually derived group-level atlas.*

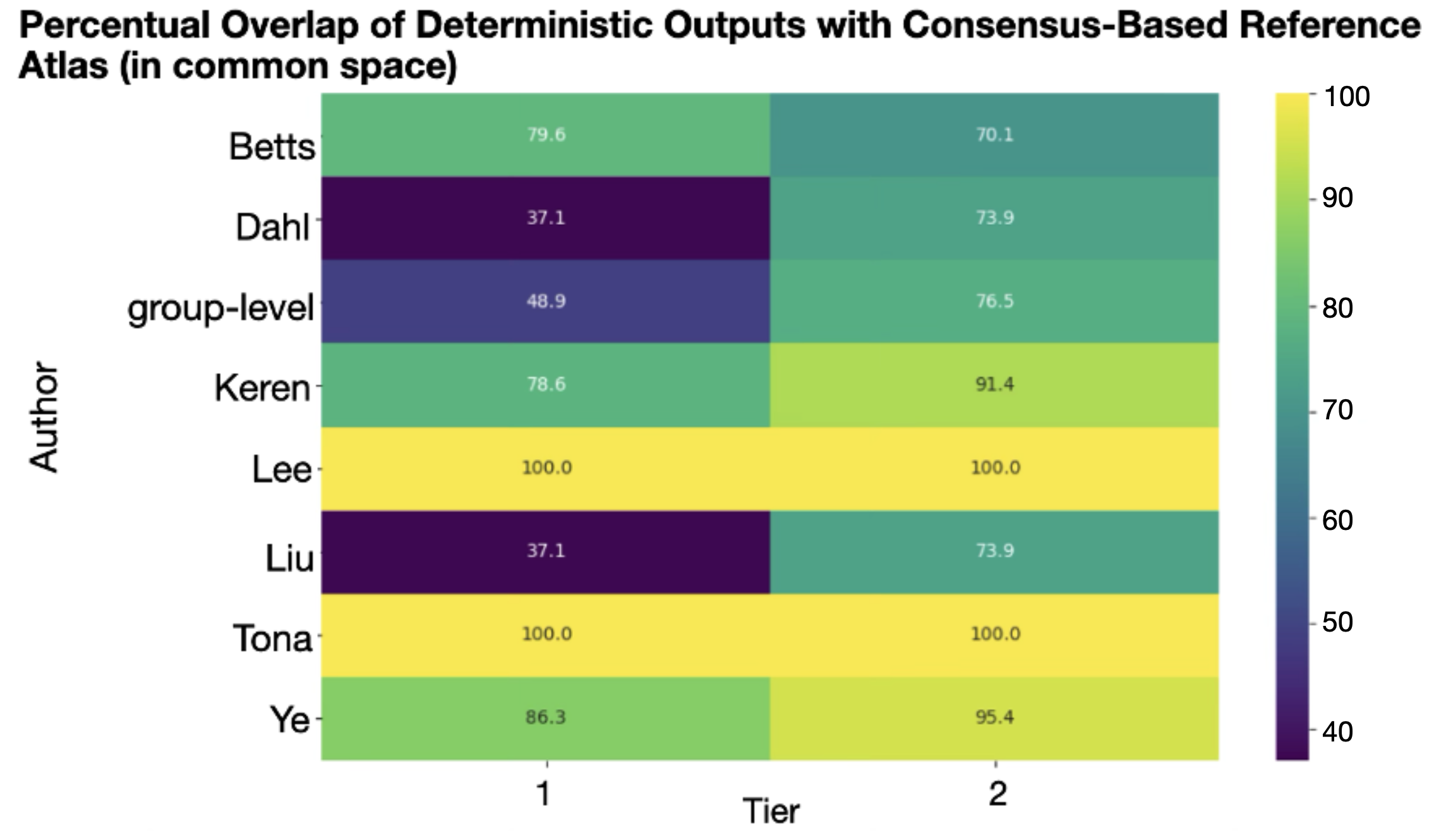

**Supplemental Figure 6**. Overlap (in %) of the atlas-based parcellation with the consensus reference atlas in MNI space (2 tier division of the LC).

*Abbreviations: group-level = manually derived group-level atlas.*

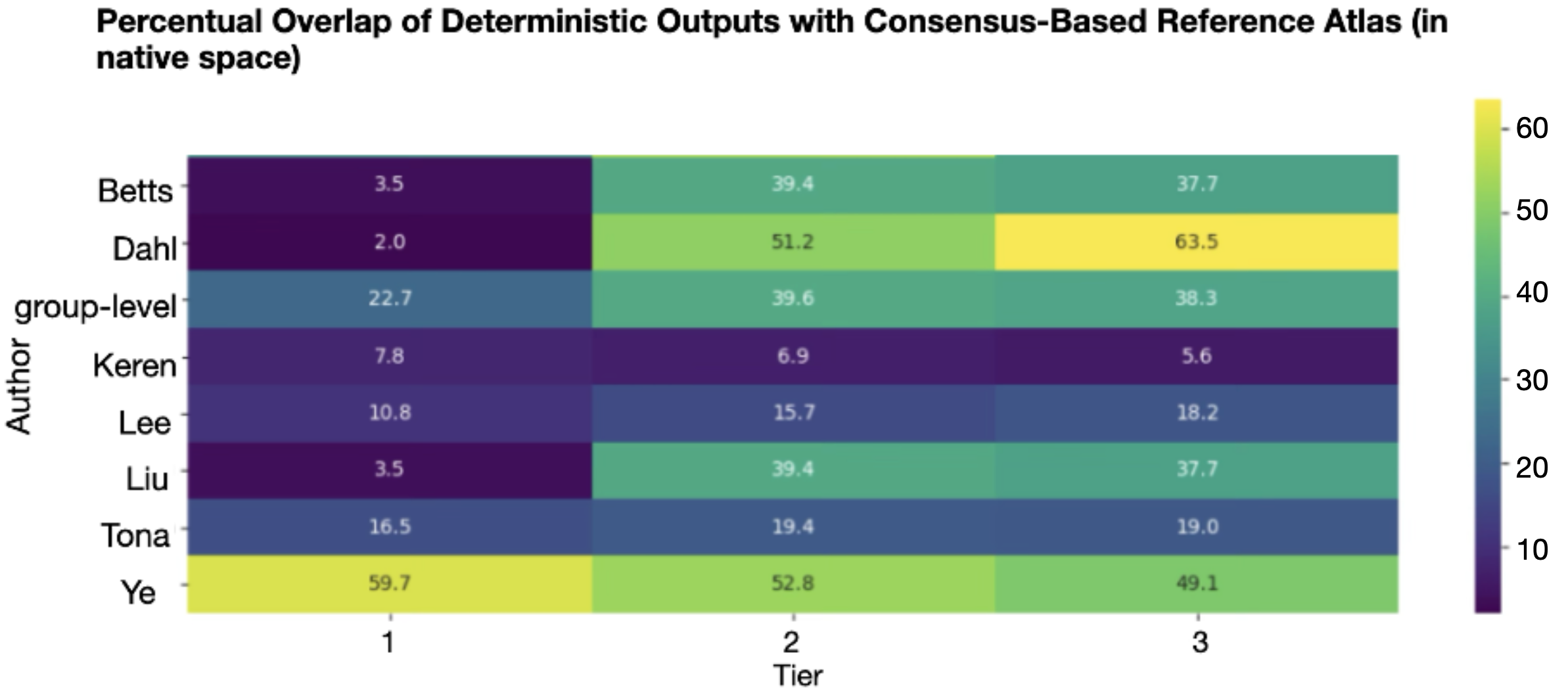

**Supplemental Figure 7**. Overlap (in %) of the atlas-based parcellation with the consensus reference atlas in native space (3 tier division of the LC).

*Abbreviations: group-level = manually derived group-level atlas.*

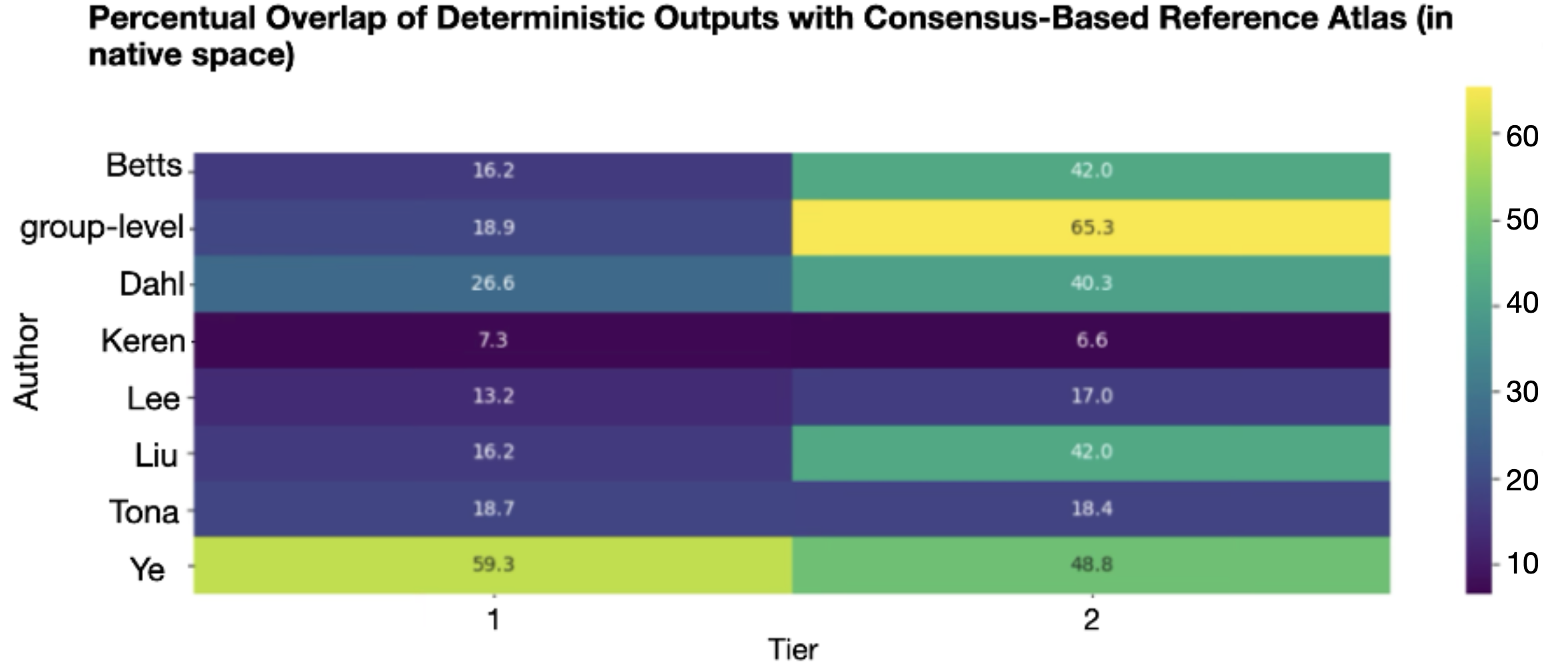

**Supplemental Figure 8.** Overlap (in %) of the atlas-based parcellation with the consensus reference atlas in native space (2 tier division of the LC).

*Abbreviations: group-level = manually derived group-level atlas.*

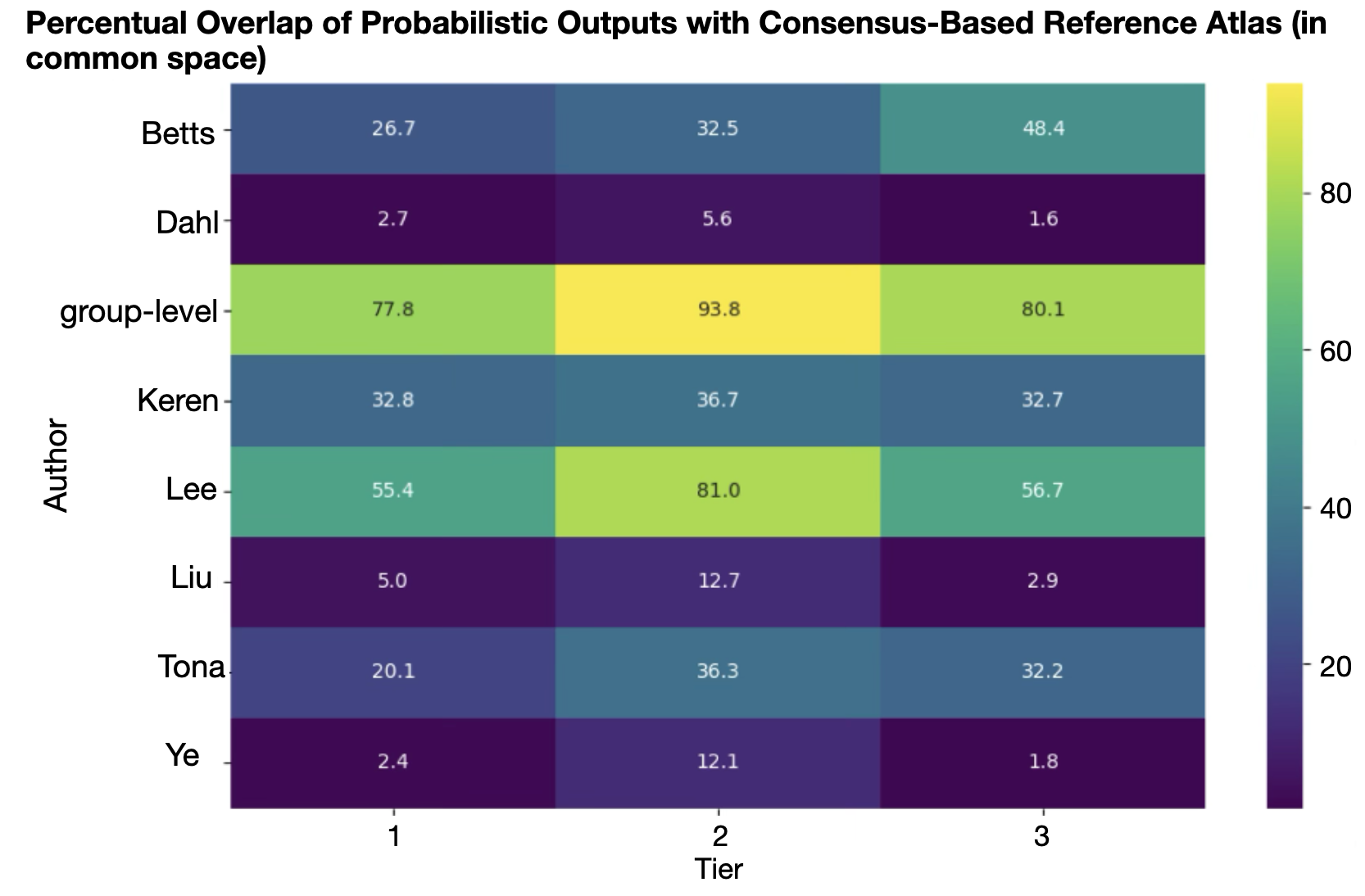
 **Supplemental Figure 9**. Overlap (in %) of the LC-enhanced tissue classification outputs with the consensus reference atlas in MNI space (3 tier division of the LC).

*Abbreviations: group-level = manually derived group-level atlas.*

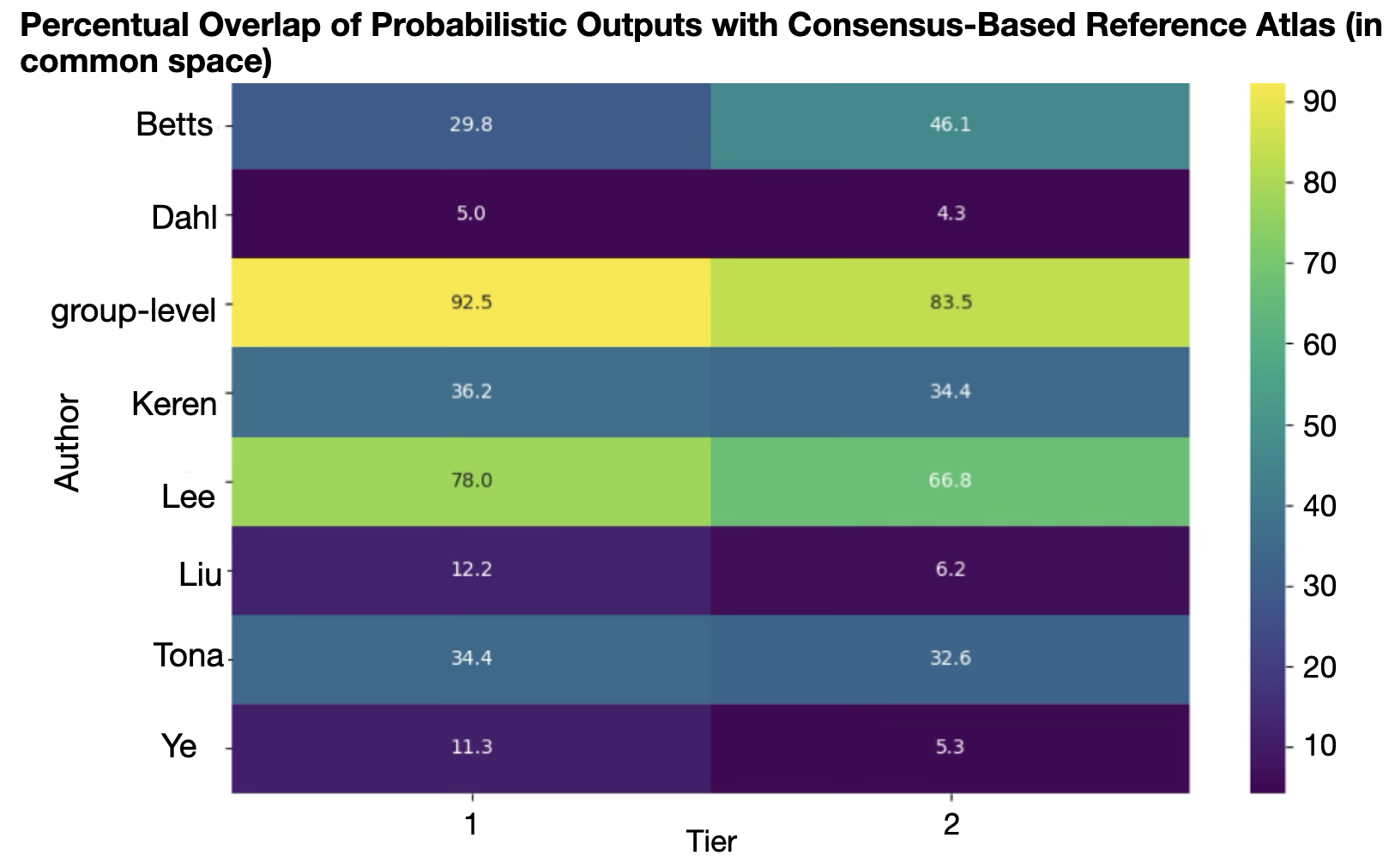

**Supplemental Figure 10**. Overlap (in %) of the LC-enhanced tissue classification outputs with the consensus reference atlas in MNI space (2 tier division of the LC).

*Abbreviations: group-level = manually derived group-level atlas.*

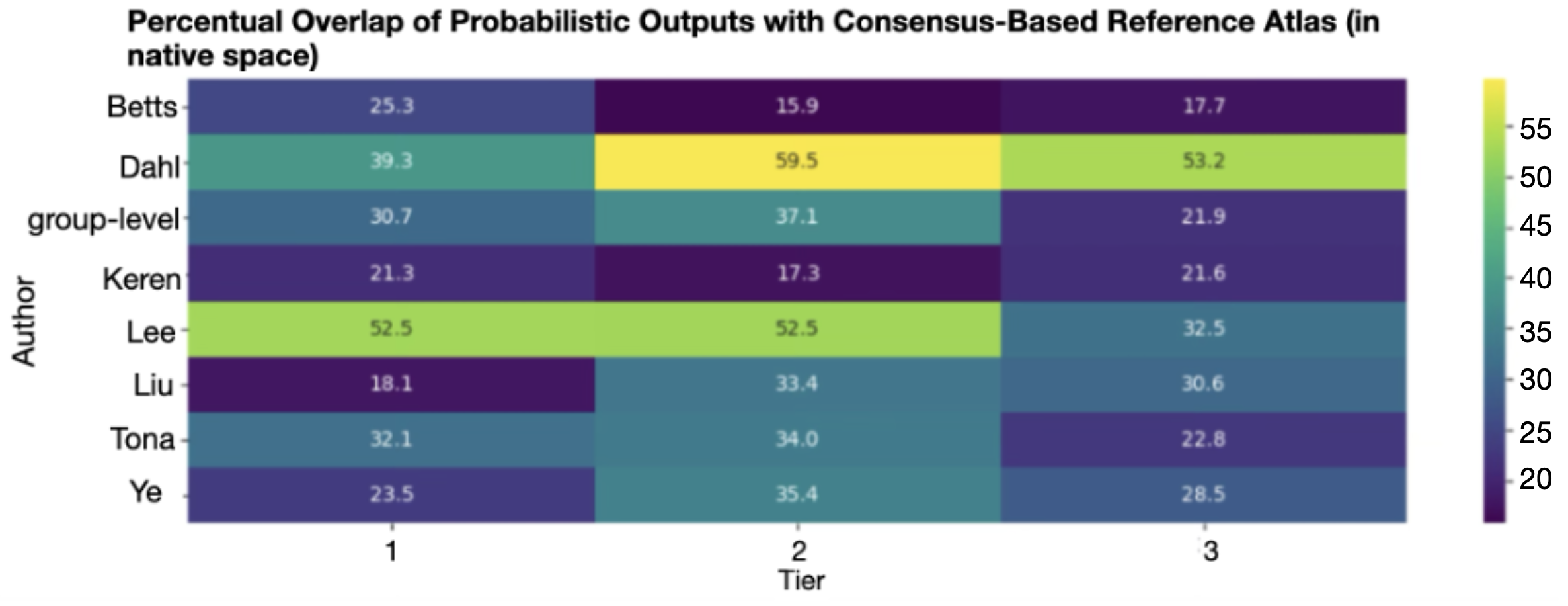
**Supplemental Figure 11**. Overlap (in %) of the LC-enhanced tissue classification outputs with the consensus reference atlas in native space (3 tier division of the LC).

*Abbreviations: group-level = manually derived group-level atlas.*

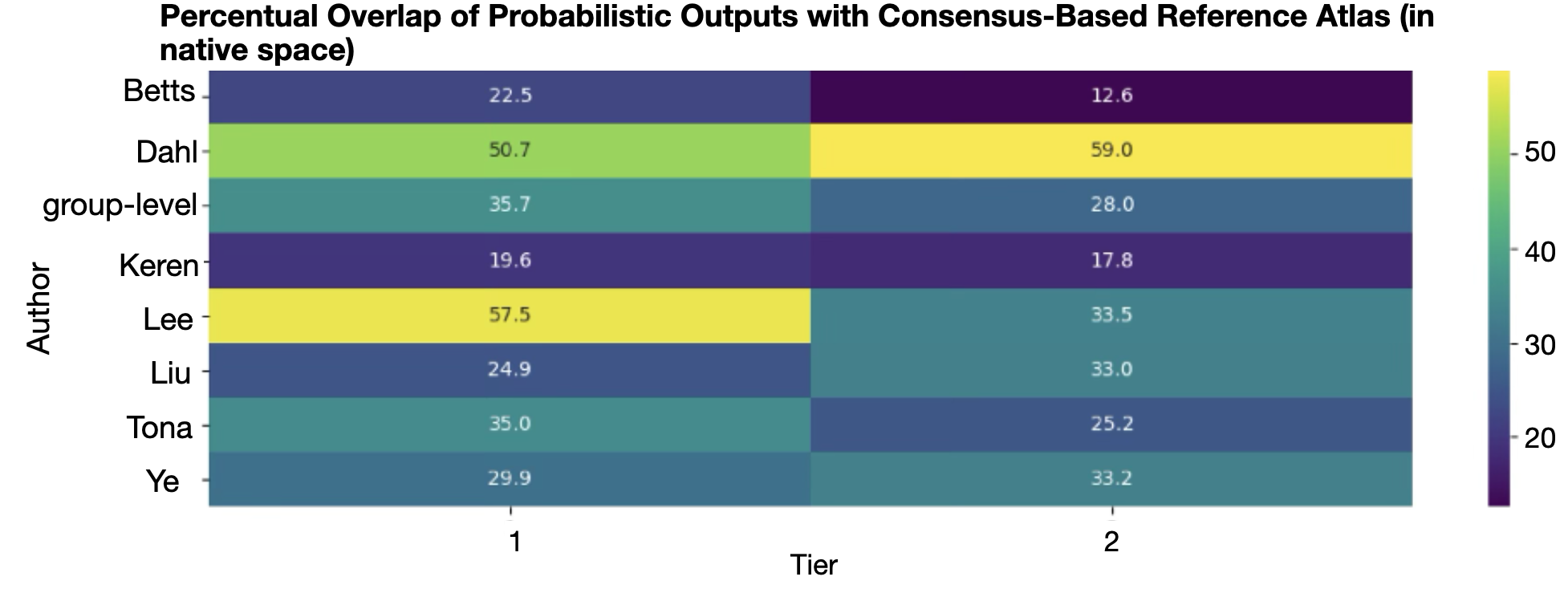

**Supplemental Figure 12**. Overlap (in %) of the LC-enhanced tissue classification outputs with the consensus reference atlas in native space (2 tier division of the LC).

*Abbreviations: group-level = manually derived group-level atlas.*

### **Tables**

**Supplemental Table 1**. Processing of atlas labels for framework.

| Author | Modifications in atlas to prepare as label in Neuromorphometrics | Modifications in atlas to prepare as 7th Tissue Class in SPM-Unified Segmentation |
| --- | --- | --- |
| Liu et al., 2019 | Spatial transformation to neuromorphometric space, downsampling to 1.5mm^3^, smoothing, binarized with Otsu threshold | Downsampling to 1mm^3^, smoothing |
| Dahl et al., 2019 | Spatial transformation to neuromorphometric space, downsampling to 1.5mm^3^, binarized with Otsu threshold | Downsampling to 1mm^3^, voxel probabilities normalized to match the previous distribution (range: 0–1) |
| Ye et al., 2021 | Spatial transformation to neuromorphometric space, downsampling to 1.5mm^3^, binarized with Otsu threshold | Downsampling to 1mm^3^, voxel probabilities normalized to match the previous distribution (range: 0–1) |
| Tona et al., 2017 | Spatial transformation to neuromorphometric space, downsampling to 1.5mm^3^, left and right LC were merged into one nifti file, binarized with Otsu threshold | Downsampling to 1mm^3^, left and right LC were merged into one nifti file, voxel probabilities normalized to match the previous distribution (range: 0–1) |
| Betts et al., 2017 | Spatial transformation to neuromorphometric space, downsampling to 1.5mm^3^, binarized with Otsu threshold | Downsampling to 1mm^3^, voxel probabilities of right side (Y=2) were changed to Y=1 |
| Keren et al., 2009 | Spatial transformation to neuromorphometric space, downsampling to 1.5mm^3^, binarized with Otsu threshold | Downsampling to 1mm^3^ |
| Lee et al., 2024 | Spatial transformation to neuromorphometric space, downsampling to 1.5mm^3^, binarized with Otsu threshold | Voxel probabilities normalized to match the previous distribution (range: 0–1) |
| Group level atlas | Spatial transformation to neuromorphometric space, downsampling to 1.5mm^3^, binarized with Otsu threshold | /. |

**Supplemental Table 2.** Comparison of automated labelling methods against manual references. Metrics include Dice Score, Volume Similarity, Cohen’s Kappa, and Hausdorff Distance. Values are reported as mean ± SD with 95% confidence intervals, rounded to two decimals.

*Abbreviations: group-level = manually derived group-level atlas.*

| Author | Metric | Mean | Median | SD | CI95 |
| --- | --- | --- | --- | --- | --- |
| group-level | Dice Score | 0.28 | 0.28 | 0.09 | 0.04 |
| Dahl | Dice Score | 0.31 | 0.32 | 0.10 | 0.04 |
| Eckert | Dice Score | 0.29 | 0.30 | 0.10 | 0.04 |
| Ye | Dice Score | 0.38 | 0.41 | 0.11 | 0.04 |
| Betts | Dice Score | 0.25 | 0.27 | 0.09 | 0.03 |
| Liu | Dice Score | 0.31 | 0.32 | 0.10 | 0.04 |
| Lee | Dice Score | 0.22 | 0.23 | 0.07 | 0.03 |
| Tona | Dice Score | 0.25 | 0.27 | 0.09 | 0.03 |
| group-level | Volume Similarity | 0.51 | 0.49 | 0.11 | 0.04 |
| Dahl | Volume Similarity | 0.58 | 0.59 | 0.11 | 0.05 |
| Eckert | Volume Similarity | 0.88 | 0.90 | 0.07 | 0.03 |
| Ye | Volume Similarity | 0.89 | 0.92 | 0.09 | 0.03 |
| Betts | Volume Similarity | 0.76 | 0.79 | 0.12 | 0.05 |
| Liu | Volume Similarity | 0.58 | 0.59 | 0.11 | 0.05 |
| Lee | Volume Similarity | 0.41 | 0.40 | 0.09 | 0.04 |
| Tona | Volume Similarity | 0.83 | 0.83 | 0.10 | 0.03 |
| group-level | Cohen’s Kappa | 0.28 | 0.28 | 0.09 | 0.04 |
| Dahl | Cohen’s Kappa | 0.31 | 0.32 | 0.10 | 0.04 |
| Eckert | Cohen’s Kappa | 0.29 | 0.30 | 0.10 | 0.04 |
| Ye | Cohen’s Kappa | 0.38 | 0.41 | 0.11 | 0.04 |
| Betts | Cohen’s Kappa | 0.25 | 0.27 | 0.09 | 0.03 |
| Liu | Cohen’s Kappa | 0.31 | 0.32 | 0.10 | 0.04 |
| Lee | Cohen’s Kappa | 0.22 | 0.23 | 0.07 | 0.03 |
| Tona | Cohen’s Kappa | 0.25 | 0.27 | 0.09 | 0.03 |
| group-level | Hausdorff Distance | 7.94 | 8.12 | 1.84 | 0.74 |
| Dahl | Hausdorff Distance | 8.36 | 8.09 | 1.55 | 0.62 |
| Eckert | Hausdorff Distance | 5.36 | 5.00 | 1.36 | 0.54 |
| Ye | Hausdorff Distance | 5.52 | 5.15 | 1.76 | 0.71 |
| Betts | Hausdorff Distance | 8.09 | 8.12 | 2.22 | 0.89 |
| Liu | Hausdorff Distance | 8.36 | 8.09 | 1.55 | 0.62 |
| Lee | Hausdorff Distance | 8.46 | 8.57 | 2.17 | 0.87 |
| Tona | Hausdorff Distance | 8.44 | 8.12 | 1.15 | 0.32 |

**Supplemental Table 3.** Comparison of automated tissue classification performance using different atlas-based spatial priors against manual labels in native space, evaluated with six metrics: Soft Dice, Volume Intersection, Voxelwise Correlation, COM Distance, KL Divergence, and EMD. Across metrics, Dahl and the group-level model generally performed best, while Keren showed lower scores. Values are reported as mean ± SD with 95% CIs, rounded to two decimals.

*Abbreviations: COM = center of mass; EMD = Earth mover’s distance; group-level = manually derived group-level atlas; KL divergence = Kullback–Leibler divergence.*

| Author | Metric | Mean | Median | SD | CI95 |
| --- | --- | --- | --- | --- | --- |
| Tona | Soft Dice | 0.14 | 0.14 | 0.09 | 0.04 |
| Ye | Soft Dice | 0.37 | 0.38 | 0.16 | 0.06 |
| Lee | Soft Dice | 0.30 | 0.30 | 0.12 | 0.05 |
| Liu | Soft Dice | 0.28 | 0.30 | 0.16 | 0.06 |
| Keren | Soft Dice | 0.11 | 0.09 | 0.08 | 0.03 |
| Dahl | Soft Dice | 0.46 | 0.47 | 0.14 | 0.06 |
| Betts | Soft Dice | 0.15 | 0.15 | 0.08 | 0.03 |
| group-level | Soft Dice | 0.42 | 0.42 | 0.19 | 0.08 |
| Tona | Volume Intersection | 16.78 | 19.43 | 9.12 | 3.73 |
| Ye | Volume Intersection | 62.64 | 59.84 | 28.92 | 11.82 |
| Lee | Volume Intersection | 41.09 | 40.87 | 17.17 | 7.02 |
| Liu | Volume Intersection | 38.00 | 45.09 | 21.40 | 8.74 |
| Keren | Volume Intersection | 13.89 | 13.92 | 10.02 | 4.10 |
| Dahl | Volume Intersection | 90.57 | 89.08 | 31.31 | 12.80 |
| Betts | Volume Intersection | 19.69 | 20.07 | 11.69 | 4.78 |
| group-level | Volume Intersection | 82.02 | 79.59 | 40.62 | 16.60 |
| Tona | Voxelwise Correlation | 0.13 | 0.18 | 0.21 | 0.08 |
| Ye | Voxelwise Correlation | 0.14 | 0.12 | 0.23 | 0.09 |
| Lee | Voxelwise Correlation | 0.16 | 0.19 | 0.19 | 0.08 |
| Liu | Voxelwise Correlation | 0.05 | 0.01 | 0.28 | 0.12 |
| Keren | Voxelwise Correlation | 0.19 | 0.18 | 0.18 | 0.07 |
| Dahl | Voxelwise Correlation | 0.15 | 0.13 | 0.24 | 0.10 |
| Betts | Voxelwise Correlation | 0.06 | 0.02 | 0.25 | 0.10 |
| group-level | Voxelwise Correlation | 0.33 | 0.31 | 0.21 | 0.08 |
| Tona | COM Distance | 6.02 | 2.48 | 11.33 | 4.63 |
| Ye | COM Distance | 1.99 | 1.78 | 1.09 | 0.45 |
| Lee | COM Distance | 2.90 | 2.68 | 1.13 | 0.46 |
| Liu | COM Distance | 5.13 | 1.51 | 11.54 | 4.72 |
| Keren | COM Distance | 9.63 | 3.99 | 13.53 | 5.53 |
| Dahl | COM Distance | 1.67 | 1.58 | 1.08 | 0.44 |
| Betts | COM Distance | 6.47 | 4.39 | 8.58 | 3.50 |
| group-level | COM Distance | 1.96 | 1.97 | 0.97 | 0.40 |
| Tona | KL Divergence | 9.86 | 9.65 | 3.08 | 1.26 |
| Ye | KL Divergence | 9.51 | 9.03 | 2.57 | 1.05 |
| Lee | KL Divergence | 8.65 | 8.67 | 2.28 | 0.93 |
| Liu | KL Divergence | 11.43 | 11.06 | 3.10 | 1.27 |
| Keren | KL Divergence | 14.72 | 14.69 | 1.82 | 0.74 |
| Dahl | KL Divergence | 7.42 | 7.64 | 2.58 | 1.06 |
| Betts | KL Divergence | 13.97 | 13.83 | 1.67 | 0.68 |
| group-level | KL Divergence | 3.91 | 3.59 | 2.47 | 1.01 |
| Tona | EMD | 1.09E-07 | 1.05E-07 | 2.25E-08 | 9.20E-09 |
| Ye | EMD | 6.77E-08 | 5.66E-08 | 2.86E-08 | 1.17E-08 |
| Lee | EMD | 6.86E-08 | 5.82E-08 | 2.07E-08 | 8.44E-09 |
| Liu | EMD | 1.07E-07 | 1.03E-07 | 2.35E-08 | 9.60E-09 |
| Keren | EMD | 1.29E-07 | 1.24E-07 | 2.10E-08 | 8.57E-09 |
| Dahl | EMD | 3.88E-08 | 3.21E-08 | 1.88E-08 | 7.67E-09 |
| Betts | EMD | 1.24E-07 | 1.23E-07 | 1.67E-08 | 6.83E-09 |
| group-level | EMD | 6.83E-08 | 7.00E-08 | 1.71E-08 | 6.98E-09 |
